# Locus coeruleus integrity is related to tau burden and memory loss in autosomal-dominant Alzheimer’s disease

**DOI:** 10.1101/2020.11.16.20232561

**Authors:** Martin J. Dahl, Mara Mather, Markus Werkle-Bergner, Briana L. Kennedy, Samuel Guzman, Kyle Hurth, Carol A. Miller, Yuchuan Qiao, Yonggang Shi, Helena C. Chui, John M. Ringman

**Author notes:** Corresponding author: MJD.

## Abstract

Abnormally phosphorylated tau, an indicator of Alzheimer’s disease, accumulates in the first decades of life in the locus coeruleus (LC), the brain’s main noradrenaline supply. However, technical challenges in reliable in-vivo assessments have impeded research into the role of the LC in Alzheimer’s disease.

We studied participants with or known to be at-risk for mutations in genes causing autosomal-dominant Alzheimer’s disease (ADAD) of early onset, providing a unique window into the pathogenesis of Alzheimer’s largely disentangled from age-related factors. Using high- resolution MRI and tau PET, we revealed lower rostral LC integrity in symptomatic participants. LC integrity was associated with individual differences in tau burden and memory decline. Post- mortem analyses in a separate set of carriers of the same mutation confirmed substantial neuronal loss in the LC.

Our findings link LC degeneration to tau burden and memory in Alzheimer’s and highlight a role of the noradrenergic system in this neurodegenerative disease.

## 1. Introduction

Alzheimer’s disease (AD) is presently an incurable neurodegenerative disease leading to dementia with the number of worldwide cases predicted to triple over the next thirty years (Alzheimer’s Disease International, 2019; Canter et al., 2016; World Health Organization, 2004). Post-mortem studies indicate that abnormally phosphorylated tau, an indicator of AD, begins to appear early in life in the locus coeruleus (LC), the primary source of cortical noradrenaline (Braak et al., 2011; Ehrenberg et al., 2017; Poe et al., 2020; Stratmann et al., 2016; Theofilas et al., 2015). With increasing age, abnormal tau appears in a characteristic topographical sequence in noradrenergic projection targets like the mediotemporal lobe (referred to as Braak stages, a classification system of the progression of tau spread; Braak et al., 2011; Chalermpalanupap, Weinshenker, & Rorabaugh, 2017; Stratmann et al., 2016), suggesting pathology slowly spreads from the LC to its target regions (Braak & Del Tredici, 2016). Consistent with this possibility, using an animal model, Gosh and colleagues demonstrated that injecting hyperphosphorylated human tau into the rodent LC leads to its slow spread to other brainstem nuclei and eventually to cortical regions (Ghosh et al., 2019).

Tau deposition is strongly linked to both neural and cognitive decline in Alzheimer’s (Hanseeuw et al., 2019; Jagust, 2018; La Joie et al., 2020). Tau pathology in the LC increases linearly with the progression of Braak stages while noradrenergic neurons first decrease in size and then degenerate (Ehrenberg et al., 2017; Kelly et al., 2017; Theofilas et al., 2017).

Indeed, a meta-analysis of twenty-four post-mortem investigations revealed substantial cell loss in the LC of Alzheimer’s patients relative to controls (mean d = 2.28; 95% CI = 2.06– 2.51; Lyness, Zarow, & Chui, 2003), and reduced neuron counts are evident already at prodromal stages of the disease (Arendt et al., 2015; Kelly et al., 2017). Topographically, noradrenergic neurodegeneration is most pronounced in rostral and middle segments of the nucleus that project to the hippocampus and other memory-relevant areas (Ehrenberg et al., 2017; Lyness et al., 2003). LC degeneration, however, does not only constitute a consequence of Alzheimer’s pathology but also may contribute to its disease development (Marien et al., 2004; Mather & Harley, 2016; Satoh & Iijima, 2019; Weinshenker, 2018). Studies with genetically modified animals indicate that abnormally phosphorylated tau in the LC leads to dysfunctional noradrenergic neuromodulation in memory-relevant brain areas (Ghosh et al., 2019; Rorabaugh et al., 2017; Weinshenker, 2018) and that tau pathology and LC degeneration synergistically aggravate neural and behavioral deterioration (Chalermpalanupap et al., 2018; also see Jacobs, Riphagen, Ramakers, & Verhey, 2019). Specifically, experimentally decreased noradrenaline levels have been associated with increased tau and amyloid-β deposition, another hallmark of AD (Chalermpalanupap et al., 2018; Heneka et al., 2010). Insights into the role of noradrenergic neurodegeneration in the progression of AD are thus of high clinical significance (Grinberg & Heinsen, 2017), yet *in-vivo* human data are scarce.

Studies in humans have been impeded by methodological challenges in non-invasive assessments of the LC due to its small size and its location deep within the brainstem (for discussions, see Astafiev, Snyder, Shulman, & Corbetta, 2010; Keren, Lozar, Harris, Morgan, & Eckert, 2009). Advances in high-resolution brainstem magnetic resonance imaging (MRI) may help the field overcome these hurdles (Betts, Kirilina, et al., 2019; Liu et al., 2017; Sun et al., 2020). Over the last fifteen years, several MRI sequences have been developed that reveal the LC as a hyperintense cluster of voxels bordering the fourth ventricle (e.g., Betts, Cardenas-Blanco, Kanowski, Jessen, & Düzel, 2017; Nakane, Nihashi, Kawai, & Naganawa, 2008; Priovoulos et al., 2017; Sasaki et al., 2006). Keren and colleagues (2015) validated a MR-based LC imaging sequence by first scanning human post-mortem samples at ultra-high field strength and afterwards performing histological analyses of the same samples. They demonstrated that the hyperintensities observed on MRI scans closely co-localize with noradrenergic cells as identified by tyrosine hydroxylase (the rate-limiting enzyme of catecholamine synthesis) staining, opening the door for non-invasive LC assessments (Keren et al., 2015; also see Cassidy et al., 2019 for a postmortem validation in dopaminergic structures and Betts, Kirilina, et al., 2019 for a discussion of potential contrast mechanisms).

Probing the utility of LC MRI to detect pathology-related changes in the noradrenergic system *in vivo*, several studies compared patients diagnosed with AD to healthy controls (Betts, Cardenas-Blanco, et al., 2019; Dordevic et al., 2017; Miyoshi et al., 2013; Takahashi et al., 2015). In general agreement with earlier post-mortem investigations (Lyness et al., 2003), the majority of studies observed lower LC MR-intensity, a proxy for the density of noradrenergic cells (Keren et al., 2015), in the patient groups (but see Miyoshi et al., 2013).

However, all of these studies investigated the by far most common, late-onset type of AD (>65 years; LOAD). LOAD is considered a heterogeneous, age-related disease that encompasses not only tau and amyloid pathology but also the aggregation of other proteins such as α-synuclein, the effects of cerebrovascular ischemia, and other processes (Jagust, 2018; Van Cauwenberghe et al., 2016). Given that LOAD incidence increases exponentially with age and so is highly correlated with age (Brookmeyer et al., 2007) and large lifespan studies suggest spatially confined age differences in LC integrity in healthy older adults (Dahl et al., 2019; Liu et al., 2019), unambiguously distinguishing disease- from age-related changes on this basis is challenging (Lindenberger et al., 2011). Focusing instead on forms of AD that begin early in life allows the possibility of distinguishing age- from disease-related changes in LC integrity (cf. Jacobs, Becker, et al., 2019).

Autosomal dominant Alzheimer’s (ADAD) is a rare inherited disease caused by mutations in genes associated with the generation or accumulation of amyloid-β (Bertram et al., 2010; Tanzi & Bertram, 2005; Van Cauwenberghe et al., 2016). In contrast to LOAD, it constitutes a more “pure” tau and amyloid pathology in which cerebrovascular- or other age-related neuropathological changes are less frequent (Jagust, 2018; Ringman et al., 2016; Spina et al., 2021) (but see Lee et al., 2016). In ADAD, symptoms develop early in life with a relatively predictable age at onset (Ryman et al., 2014), thus providing a unique window into the pathogenesis of Alzheimer’s (Bateman et al., 2012; Ringman, 2005). Whether or not there is MR-identifiable LC degeneration in patients diagnosed with Alzheimer’s dementia early in life is still an open question. Importantly, answering this question could help shed light on whether LC involvement is a core feature of AD or whether it should be considered more indicative of a non-Alzheimer’s process generally associated with aging, such as the process outlined in the “primary age-related tauopathy” framework (Crary et al., 2014).

While previous post-mortem research suggests a topographical pattern of noradrenergic neurodegeneration within the LC in LOAD (Ehrenberg et al., 2017; Lyness et al., 2003), most current *in-vivo* (MRI) studies lack the spatial specificity to draw comparable conclusions. That is, they report disease-related differences in MR-indexed LC integrity (Dordevic et al., 2017; Takahashi et al., 2015) without exploring potential topographical patterns therein, precluding direct comparisons (but see Betts, Cardenas-Blanco, et al., 2019). Investigations in healthy participants successfully applied LC MRI to map the spatial extent of the central noradrenergic nucleus in standard space (Betts et al., 2017; Dahl et al., 2019; Keren et al., 2009; Liu et al., 2019; Tona et al., 2017; Ye et al., 2021), paving the way for topographical cross-study comparisons. However, estimates of locus coeruleus’ dimensions and localizations show a sizeable variance across publications (range of agreement: 48–94%; cf. Dahl et al., 2019; Liu et al., 2019; Ye et al., 2021; or even 1–40% using a different methodology, Mäki-Marttunen & Espeseth, 2020). Such lack of consensus considerably limits the replicability of findings and impedes scientific progress.

Thus, the present study pursued two main goals: First, we aimed at improving the reliability and validity of MR-based LC detection. Leveraging a meta-analytical approach, we aggregated across previously published maps of the LC (Betts et al., 2017; Dahl et al., 2019; Keren et al., 2009; Liu et al., 2019; Tona et al., 2017; Ye et al., 2021) to derive a biologically plausible volume of interest (meta mask) that shows high agreement across investigations.

Second, we aimed at determining whether MR-indexed LC integrity can serve as marker for noradrenergic degeneration in early-onset ADAD. To this end, we applied the newly generated meta mask to an independent clinical sample to semi-automatically extract MR- indexed LC integrity across the rostrocaudal extent of the nucleus (cf. Dahl et al., 2019). We specifically focused on a sample of participants with or known to be at-risk for rare mutations in genes causing ADAD (Presenilin-1 protein gene [*PSEN1*]; Amyloid-β Precursor Protein gene ; Goate et al., 1991; Murrell et al., 2006; Yescas et al., 2006). In addition, using an independent sample of banked post-mortem neuropathological specimens, we characterized changes occurring in the LC in persons dying with the A431E mutation in *PSEN1* relative to controls. We hypothesized that middle to rostral MR-indexed LC integrity would be lower in living symptomatic participants relative to healthy matched controls, corroborating earlier post-mortem findings (in LOAD; Lyness et al., 2003). Similarly, post-mortem LC specimens were hypothesized to show signs of neurodegeneration in ADAD relative to controls. Finally, we predicted MR-indexed LC integrity would be associated with cortical tau burden (Chalermpalanupap et al., 2018), as assessed using positron emission tomography (PET), and attention and memory performance (Dahl et al., 2019; Elman et al., 2021; Liu et al., 2020).

## 2. Material and methods

### 2.1.1. Aggregating across published locus coeruleus maps to derive a high confidence meta mask

Previous mapping studies used a variety of LC-sensitive MRI sequences (Turbo Spin Echo [TSE; Dahl et al., 2019; Keren et al., 2009; Tona et al., 2017]; Magnetization Transfer [MT; Liu et al., 2019; Ye et al., 2021]; Fast Low Angle Shot [FLASH; Betts et al., 2017]) at different field strength (7 Tesla: Ye et al., 2021; 3 Tesla: all other studies) to visualize the nucleus. Most investigations were conducted across younger and older adults or in lifespan samples (but see Tona et al., 2017 and Ye et al., 2021 for studies in younger and older adults, respectively). After data collection, individual brainstem scans were transformed into a group or standard space in which a manual (Betts et al., 2017; Liu et al., 2019; Tona et al., 2017) or semi-automatic (Dahl et al., 2019; Keren et al., 2009; Ye et al., 2021) approach was taken to segment the LC from surrounding tissue. While some earlier reports (Dahl et al., 2019; Liu et al., 2019; Ye et al., 2021) include a comparison of a subset of the published masks, to date there is no systematic evaluation of their agreement. More importantly, no previous study attempted to resolve the apparent disagreement in LC dimensions and localizations across publications.

In keeping with earlier analyses (Dahl et al., 2019; Liu et al., 2019; Ye et al., 2021), the LC masks noted in Table 1 were used for cross-study comparisons. Before relating different masks to one-another, all volumes were binarized and moved to a common space (MNI-ICBM 152 linear space, 0.5 mm resolution), if necessary. In standard space, first an unthresholded aggregate mask was generated using the following formula:

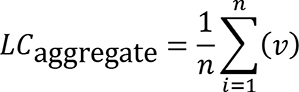

**Table 1:** Locus coeruleus masks included in comparisons

| Publication | MRI acquisition | Sample | Locus coeruleus segmentaion | Mask source | Mask version |
| --- | --- | --- | --- | --- | --- |
| Betts et al., 2017 | 3 T; FLASH | N = 82; YA; OA | Manual | https://doi.org/10.1016/j.neuroimage.2017.09.042 | -- |
| Dahl et al., 2019 | 3 T; TSE | N = 294; YA; OA | Semi-automatic | https://www.mpib-berlin.mpg.de/LC-Map | -- |
| Keren et al., 2009 | 3 T; TSE | N = 44; YA–OA | Semi-automatic | http://www.eckertlab.org/LC | 1 SD |
| Liu et al., 2019 | 3 T; MT | N = 605; YA–OA | Manual | https://osf.io/r2bwk | -- |
| Tona et al. 2017 | 3 T; TSE | N = 17; YA | Manual | http://www.nitrc.org/projects/prob_lc_3t | <sup>1</sup> |
| Ye et al., 2021 | 7 T; MT | N = 53; OA | Semi-automatic | https://www.nitrc.org/projects/lc_7t_prob | 5% prob.<sup>2</sup> |

Note: <sup>1</sup> The masks provided by Tona and colleagues were averaged across scan sessions and segmentations. <sup>2</sup> The mask by Ye and colleagues was linearly and non-linearly co-registered from MNI-ICBM 152 non-linear 2009b to MNI-ICBM 152 linear space (0.5 mm resolution) using Advanced Normalization Tools (antsRegistration). MNI-ICBM 152 linear was chosen as target space as all but one masks share this space. For co-registration, the standard brains of the respective spaces were aligned and the transformations were applied to the mask file with a nearest neighbor interpolation. The transformations from MNI 2009b non-lin ↔ MNI linear are shared via osf.io/sf2ky/; T, Tesla; FLASH, Fast Long Angle Shot; TSE, Turbo Spin Echo; MT, Magnetization Transfer; YA, Younger Adults; OA, Older Adults; YA–OA, lifespan sample including younger, middle-aged and older adults; SD, standard deviation; prob., probability.

Whereby *n* denotes the number of masks included in the analyses (*n* = 6) and *v* denotes the value for a given voxel (either zero [non-LC tissue] or one [LC]). That is, at every voxel the individual binarized masks were averaged, resulting in an aggregate mask (*LCaggregate*; see Figure 1). The generated volume, *LCaggregate*, showed a value range between 0 and 1 (complete agreement that a voxel does not/does belong to the LC, respectively) in steps of ^1^. Please note that we decided against weighing the contribution of the individual masks to the *LCaggregate* by sample size as two masks (Dahl et al., 2019; Liu et al., 2019) account for more than 80% of the total number of participants (see Table 1). Hence, the aggregate would be strongly biased in favor of a subset of masks that are themselves highly spatially congruent (agreement 94%; Liu et al., 2019).

**Figure 1.**
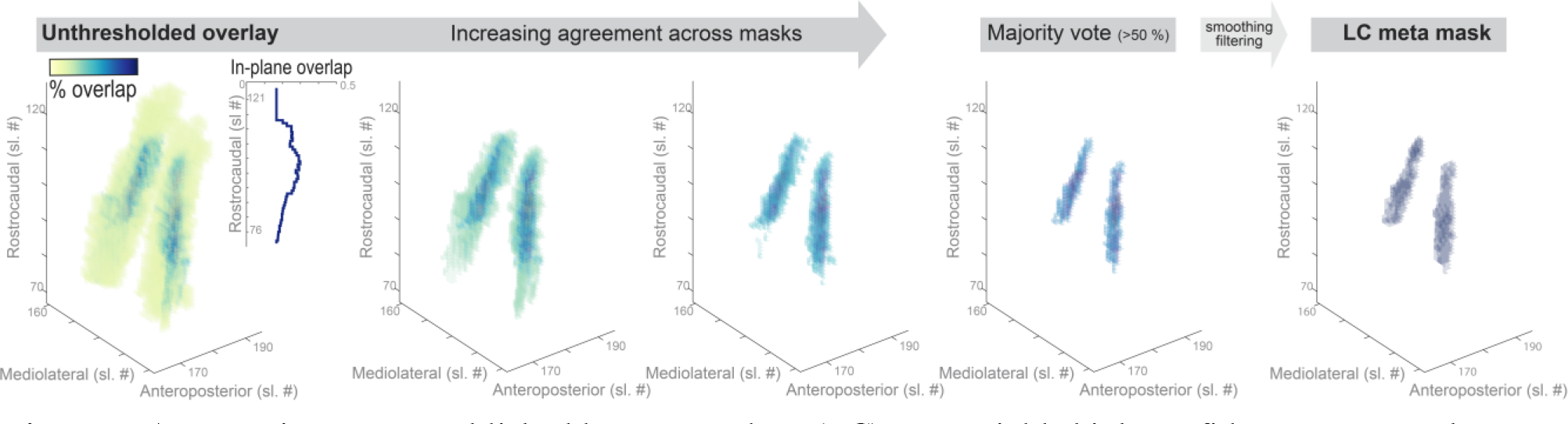
Aggregating across published locus coeruleus (LC) maps yields high confidence meta mask. **Left**. Unthresholded voxel-by-voxel average across published LC masks. Areas with voxels that are consistently judged to belong to the LC are shown in dark blue while low agreement areas are shown in yellow. **Inlay**. Agreement across masks (averaged across voxels in-plane) is lowest in most rostral and caudal regions. **Middle**. Pruning low agreement areas isolates a core of high confidence voxels. **Right**. Smoothing and filtering the thresholded overlay yields a biologically plausible LC volume of interest (rostrocaudal extent of 14 mm; volume of 74 mm³ [592 voxels]), corresponding well with earlier post- mortem reports (Fernandes et al., 2012). Sl. #, slice number in MNI 152 lin (0.5 mm) space; The LC meta mask is available for download via: osf.io/sf2ky/).

In a second step, the aggregate mask was pruned by removing low-agreement voxels until a majority vote was achieved. That is, only areas with agreement values > 50% were retained (i.e., more than ½*n* masks voted for a voxel to belong to the LC). Essentially, the described mask generation process is comparable to label fusion with majority voting (Rohlfing et al., 2004; Sabuncu et al., 2010; Wang et al., 2013).

Finally, to obtain a biologically more plausible volume, the pruned aggregate mask was smoothed with a 0.1 mm full width at half maximum (FWHM) kernel using SPM12 (Penny et al., 2007) in Matlab (The MathWorks Inc., Natick, MA, USA). Voxels exceeding a threshold of 0.05 were considered part of the LC in the meta mask (*LCmetaMask*; see Figure 1).

The resulting mask had a rostrocaudal extent of 14 mm and a volume of 74 mm³ (592 voxels), corresponding well with previous post-mortem findings (Fernandes et al., 2012; Tona et al., 2017). We share the generated meta mask with the scientific community via osf.io/sf2ky/

### 2.1.2 Evaluation of previously published locus coeruleus masks

Once a high-agreement volume of interest (*LCmetaMask*) was established, published locus coeruleus masks were compared regarding their specificity and sensitivity. That is, we evaluated (1) how many voxels of a given mask fell within the *LCmetaMask* relative to the total number of voxels in that mask and (2) how many voxels of the *LCmetaMask* were included in each mask (relative to the total number of voxels in the *LCmetaMask*). The mean of the quality metrics (specificity, sensitivity) was taken as indicator for the accuracy of a mask.

### 2.2. Application of locus coeruleus meta mask to an independent clinical sample

#### 2.2.1. Study design and participants

To determine whether the *LCmetaMask* can detect clinically significant differences in LC integrity, eighteen participants with or known to be at-risk for mutations in genes associated with ADAD (*PSEN1* or *APP*) were investigated (34.7 ± 10.1 years [mean ± standard deviation]; 9♀; see Table 2; data collection was interrupted by the COVID-19 pandemic).

**Table 2:** Descriptive statistics for imaging subgroups

|  | ADAD-control | ADAD-presymptomatic | ADAD-symptomatic |
| --- | --- | --- | --- |
| n | 9 | 5 | 4 |
| Mutation | 0/9 | 5/5 | 4/4 |
| Mutation type |  | *PSEN1* (A431E; 3/5)<br>*PSEN1* (I180F; 1/5)<br>*APP* (V717I; 1/4) | *PSEN1* (A431E; 3/4)<br>*APP* (V717I; 1/4) |
| Diagnosis | 0/9 | 0/5 | 4/4 |
| CASI | 95.3 | 95.0 | 74.9 |
| (std, range) | (1.8; 92–97.7) | (4.2; 90–99) | (11.8; 62.8–86.8) |
| CDR | 0 | 0 | 0.75 |
| (std, range) | (0, 0) | (0, 0) | (0.3; 0.5–1) |
| Age | 33.4 | 30 | 43.5 |
| (std, range) | (12.2; 19–58) | (4.5; 22–33) | (1.3; 42–45) |
| Gender | 5 male<br>4 female | 1 male<br>4 female | 3 male<br>1 female |

*Note:* ADAD, autosomal-dominant Alzheimer's disease; ADAD-control: control participants not carrying the mutation, but genetically related to mutation carriers; ADAD-presymptomatic: presymptomatic mutation carriers (Washington University Clinical Dementia Rating Scale [CDR] scores of 0); ADAD-symptomatic: symptomatic mutation carriers; std. standard deviation; CASI, Cognitive Abilities Screening Instrument (Teng et al., 1994); *PSEN1*: Presenilin-1 protein (gene); *APP*: Amyloid-β Precursor Protein (gene)

**Table 3:** Neuropathological assessment of amyloid β deposits (A), staging of neurofibrillary tangles (B), and scoring of neuritic plaques (C)*, as well as Braak and Braak** staging for specimen of mutation carriers

| Case | A | B | C | Braak & Braak |
| --- | --- | --- | --- | --- |
| Case 1024 | 3 | 3 | 3 | VI |
| Case 412<sup>1</sup> | 3 | 3 | 3 | VI |
| Case 838<sup>2</sup> | 3 | 3 | 3 | VI |
| Case 697 | 3 | 3 | 3 | VI |

Note: <sup>1</sup> Staging performed in 1996; <sup>2</sup> No official score available for CASE 838, provided staging indicates an estimate by the pathologist.
\*(Montine et al., 2012); \*\* (Braak et al., 2011)

Participants’ consent was obtained according to the Declaration of Helsinki. The study was approved by the ethical committee of the University of Southern California. Data were acquired as part of a study employing the Human Connectome Protocol (HCP) in genetic subtypes of AD (NIH U01AG051218, PI: Ringman; for more details, see adrc.usc.edu/scientists-researchers/ [Estudio de la enfermaded de Alzheimer en Jalisciences; EEAJ]). Genotyping confirmed mutations in nine of the eighteen participants (the A431E substitution in *PSEN1*, n = 6; the I180F substitution in *PSEN1*, n = 1; and the V717I substitution in *APP*, n = 2; Goate et al., 1991; Murrell et al., 2006; Yescas et al., 2006), of which four were symptomatic. We defined symptomatology based on the Washington University Clinical Dementia Rating Scale [CDR] (see Table 2). Symptomatic participants showed CDR values greater than zero, while all participants with a value of zero were considered cognitively normal (n = 14).

Participants underwent 3 T MRI, flortaucipir PET, and cognitive testing. LC MR-intensity, a non-invasive proxy for neuronal density (Keren et al., 2015), was semi-automatically extracted from high-resolution brainstem scans (Dahl et al., 2019). LC ratios— that is, a ratio of peak LC intensity standardized to a pontine reference (Betts, Kirilina, et al., 2019; Liu et al., 2017)—were computed across the rostrocaudal extent of the nucleus.

Standard uptake value ratios (SUVR) were calculated from partial volume corrected flortaucipir PET data using cerebellar gray matter as the reference based on the PETSurfer tool from FreeSurfer (Greve et al., 2014). SUVR images were mapped to the cortical surface which was parcellated into 36 regions of interest (Desikan et al., 2006). Univariate associations between LC ratios, clinical variables, cognitive status, and tau burden were evaluated using non-parametric Wilcoxon rank sum tests and bootstrapped Spearman’s correlations. For Wilcoxon rank sum tests, effect sizes (*r*) are calculated by dividing the test statistic (*Z*) by the square root of the sample size (n; (Rosenthal, 1991)). The multivariate pattern between LC ratios, cortical tau burden and cognitive decline was evaluated using a partial least squares correlation (see below; Krishnan, Williams, McIntosh, & Abdi, 2011; McIntosh & Lobaugh, 2004). For bivariate correlations and partial least squares correlations, we provide the rho values as effect size measure. Two-tailed statistical tests were used with an alpha level of *p* < 0.05. The applied statistical tests did not include covariates. Participants with missing data were excluded from the respective analyses (see below). We did not conduct a formal power calculation, given that there was no available prior evidence on the studied phenomena.

#### 2.2.2. Assessment of imaging data

Structural MRI data were collected employing a 3 T Siemens Prisma scanner with a 32-channel head coil following a Human Connectome Project imaging approach (HCP; cf. humanconnectome.org/). PET data were acquired on a 64-slice Biograph PET/CT Siemens scanner at the USC PET Imaging Sciences Center. Only those sequences used in the current analyses are described below.

A three-dimensional (3D) T1-weighted magnetization prepared gradient-echo (MPRAGE) sequence with a duration of 6:38 min and the following parameters was applied: repetition time (TR) = 2400 ms; echo time (TE) = 2.22 ms; inversion time (TI) = 1000 ms; flip angle = 8°; bandwidth = 220 Hz/Px; dimensions = 208 × 300 × 320; isometric voxel size = 0.8 mm^3^. Based on this whole-brain (MPRAGE) sequence, a high-resolution, two-dimensional (2D) T1-weighted TSE sequence was aligned perpendicularly to the plane of the brainstem. Acquisition of the TSE sequence took 1:53 min, and the following parameters were used: TR = 750 ms; TE = 10 ms; flip angle = 120°; bandwidth = 285 Hz/Px; dimensions = 512 × 512 × 11; anisometric voxel size = 0.43 × 0.43 × 3.5 mm^3^ (mimicking the elongated shape of the locus coeruleus). Each TSE scan consisted of eleven axial slices.

PET data were collected within the period of 90–120 min after the injection of the compound 18F-AV-1451 (total dose: 344 MBq). Low dose computerized tomography (CT) scans were acquired prior to the PET scans for attenuation correction. The dynamic PET scans included six frames of five-minute duration, with the dimensions of 168 × 168 × 56 and a voxel size of 1.5 × 1.5 × 4 mm^3^. The six dynamic scans were rigidly aligned to the first one and averaged to create a static image for SUVR-based analysis.

#### 2.2.3. Semi-automatic locus coeruleus intensity assessment using meta mask

Leveraging the generated *LCmetaMask* (see above), locus coeruleus intensity was semi-automatically extracted using a previously validated pipeline (Dahl et al., 2019; also see Ye et al., 2021 for a comparable approach). In short, individual whole brain and brainstem scans were iteratively aligned across participants using a template-based procedure implemented in Advanced Normalization Tools (version 2.1; ANTs; Avants et al., 2011; Avants, Tustison, & Song, 2009), and subsequently transformed to standard space (MNI-ICBM 152 linear, 0.5 mm; see Dahl et al., 2019 for a step-by-step description of the standardization). In standard space, individual brainstem scans were masked using the high-confidence *LCmetaMask* to remove non-LC tissue (using SPM12 in Matlab). To allow inter-participant comparisons of LC data, a normalization of the arbitrarily scaled MR-intensity values is required (Betts, Kirilina, et al., 2019). Thus, brainstem scans were additionally masked using a reference volume of interest positioned in the central pontine white matter, as previously suggested (Ye et al., 2021; dimensions: 4 × 4 mm in plane, following the *LCmetaMask* with a constant distance of 8.5 mm [y-dimension] along the rostrocaudal axis; see Figure 3). Within the masked brainstem scans, we then fully automatically searched for voxels of brightest intensity in the LC and reference regions. Next, individual, spatially-resolved LC intensity ratios were computed for each slice using the following formula (Dahl et al., 2019; Liu et al., 2017):

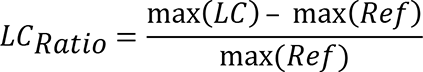

Where max(*LC*) denotes the peak intensity for a given slice in the LC volume of interest and max(*Ref*) indicates the peak intensity in the reference region. After extraction, ratios of the left and right hemisphere were averaged for further analyses to obtain more stable intensity estimates. Finally, the peak ratio across the rostrocaudal LC extent was calculated as proxy for overall locus coeruleus integrity.

#### 2.2.4. Assessment of cortical tau burden

To evaluate the association of MR-indexed LC integrity to hallmarks of AD, we calculated cortical tau burden from flortaucipir PET data.

First, each participant’s whole brain MR data (MPRAGE) was preprocessed using a HCP pipeline (version 3.27; Makropoulos et al., 2018). Next, the cortical surface was reconstructed and parcellated into thirty-six anatomical regions of interest using Freesurfer 6.0 (Dale et al., 1999; Desikan et al., 2006). Flortaucipir PET images were then co-registered within participants to whole brain (MPRAGE) native space. The Muller-Gartner (MG) method was used for partial volume correction of the PET data, as implemented in PETSurfer (Greve et al., 2016). Standard uptake value ratios (SUVR) were calculated using cerebellar gray matter as reference region. Finally, partial-volume corrected SUVR images were mapped to the cortical surface and parcellated into thirty-six regions of interest (Desikan et al., 2006). For further analyses, each region’s SUVR data were averaged across hemispheres to obtain more stable estimates of cortical tau burden. Tau PET data were not available for two participants.

#### 2.2.5. Association of locus coeruleus intensity and cortical tau burden

We employed a multivariate statistical approach to assess the relation between MR- indexed locus coeruleus integrity and tau pathology across the cortex. Specifically, using a Partial Least Squares Correlation (PLSC; Krishnan et al., 2011; McIntosh & Lobaugh, 2004) we estimated a latent tau pathology profile that is maximally related to LC integrity (cf. Keresztes et al., 2017; Muehlroth et al., 2020).

First, a between-person Pearson’s correlation matrix (*R*) was computed between the overall LC ratios (i.e., a vector *YLC* with *n*Participants × 1 [overall intensity ratios]) and the tau SUVR data (i.e., a matrix *XTau* with *n*Participants × 36 [regions of interest]). This correlation matrix (*RLC,Tau* = *YLC*^T^*XTau*) was then decomposed into three matrices using a singular value decomposition (SVD; SVD(*RLC,Tau*) = *USV*^T^). Whereby *U* refers to a left singular vector of LC weights, *V*^T^ denotes a right singular matrix of regional tau weights, and *S* is a diagonal matrix of singular values. Based on this matrix decomposition, a single latent variable was extracted. The latent variable optimally expresses (in a least squares sense) the pattern of interindividual differences in regional tau burden that shares the largest amount of variance with interindividual differences in overall LC intensity ratios. The statistical strength of the extracted pattern (i.e., the latent variable) was evaluated using a permutation test (by randomly re-ordering the observations in *XTau* while leaving *YLC* unchanged and re-calculating the SVD; *n*Permutations = 10,000). Subsequently, the reliability of the contribution (i.e., weights [*V*^T^]) of individual cortical regions, that is, the columns in *XTau*, to this latent variable was determined using a bootstrapping procedure (*n*Bootraps = 10,000). A ratio of the region-specific weights (*V*^T^) and their corresponding bootstrapped standard errors provided bootstrap ratios (BSR) that can be interpreted akin to Z-scores. Finally, multiplying participants’ regional pattern of tau pathology (*XTau*) with each regions’ contribution to the latent variable (i.e., its weight, *V*^T^) yielded a summary measure, reflecting participants’ LC-related tau pathology.

#### 2.2.6. Cognitive assessments

Participants completed a series of standardized neuropsychological tests to assess memory and attention. The cognitive assessment included screening instruments (the Montreal Cognitive Assessment [MoCA, Nasreddine et al., 2005]; the Cognitive Abilities Screening Instrument [CASI; Teng et al., 1994], and the Washington University Clinical Dementia Rating Scale [CDR; measure: sum of boxes; cf. O’Bryant et al., 2008]). In addition, attention (Digit-Symbol-Substitution Test [DSST; Wechsler, 1981]; Digit Span [measure: backward report; Wechsler, 1997], Trail Making Test [measure: part B; Tombaugh, 2004]) and memory performance were assessed (the Spanish English Verbal Learning Test [SEVLT; measures: average performance over learning trials and delayed recall; González, Mungas, & Haan, 2002], a learning and memory test akin to the Rey Auditory Verbal Learning Test [RAVLT] in which Spanish and English versions were simultaneously developed; Benson Complex Figure Test [measure: delayed recall; cf. Possin, Laluz, Alcantar, Miller, & Kramer, 2011]; and the Craft Story Test [delayed recall; Craft et al., 1996]). When available, for further analyses individual test scores were transformed to Z-scores using published normative data. Trail Making Test data were not available for one participant.

A partial least squares correlation (PLSC; Krishnan et al., 2011; McIntosh & Lobaugh, 2004) was applied to capture the multivariate association between MR-indexed LC integrity and cognitive performance, using the same methods as described above.

#### 2.3.1 Neuropathological assessment

To characterize the pathology underlying the LC signal on MRI in ADAD, we examined the LC in an independent sample of four persons dying with the A431E mutation in *PSEN1* in comparison to five age-matched neurologically normal controls. In short, paraffin-embedded sections of the pons were cut and stained with hematoxyin and eosin (H&E), glial fibrillary acid protein (GFAP), AT8 antibodies for neurofibrillary pathology, 4G8 antibodies for amyloid-β, and anti-alpha synuclein antibodies for Lewy Body pathology. Qualitative comparisons were made between patients and controls.

#### 2.3.2. Histopathology methods

After 1-month fixation, tissue areas were dissected and paraffin-embedded and serially sectioned into 5 µm-thick sections using a microtome (Microm International Waldorf, Germany). Brain sections were counterstained with H&E and reagents were purchased from Sigma-Aldrich (St. Louis, MO, USA). Sections adjacent to H&E were immunostained using the Leica Bond RXm™ automated staining processor (Leica Biosystems, Buffalo Grove, IL, USA). Tissue sections were stained using the Bond Polymer Refine AFP (Red) Detection System for all Abs except GFAP Ab and NeuN Ab which used Bond Polymer Refine DAB Detection System (Leica Biosystems). Antigen retrieval was performed with Epitope Retrieval Solution 1 (ER1, pH 6) for 20 min except for GFAP Ab which was for 10 min. Sections were then incubated with amyloid-beta (4G8, BioLegend, SIG #800701, 1:4000) for 30 min, TAR DNA binding protein (TDP-43, Sigma, T11705; 1:1250) for 15 min, phosphor-tau (AT8, MN1020, Thermo Scientific, USA; 1:1000 for 15 min; GFAP Rb pAb, Chemicon, Temecula AB5804, CA, USA; 1;2500) for 15 min; (NeuN, AbCAM, ab104224, USA; 1:800) for 30m. a- Synuclein Rb pAb, Chemicon, AB5038, 1:2000 for 15 min.

Staining with 4-repeat isoform tau (RD4, Cosmo Bio Co. LTD. Inspiration of Life Science, Cat# TIP-4RT-P01; 1:3000) and 3-repeat isoform tau (RD3, 05-803, Millipore, USA; 1:5000) required manual pretreatment outside autostainer. Sections heated in a pressure cooker at 125° in citrate buffer (CC2, pH 6) for 90 min and treated with 88% Formic Acid for 10 min. Sections were then loaded in autostainer with Bond Polymer Refine Red Detection System for 15 min. Images were obtained with a Nikon Eclipse E400 brightfield microscope (Nikon Instruments Inc, Melville, NY, USA). All stains were performed at Children’s Hospital of Los Angeles.

### 2.4. Data and code availability

The here described LC meta mask as well as pontine reference mask is available for download on an Open Science Framework repository (osf.io/sf2ky/). All sources for previously published masks are listed in Table 1. For access to raw imaging and clinical data, please refer to the HCP (https://nda.nih.gov/ccf). The customized code used for these analyses is available from the corresponding authors upon request.

### 3. Results

### 3.1.1. Aggregating across published locus coeruleus maps yields high confidence meta mask

Previously published masks of the LC (Betts et al., 2017; Dahl et al., 2019; Keren et al., 2009; Liu et al., 2019; Tona et al., 2017; Ye et al., 2021) were averaged voxel-per-voxel to generate an unthresholded overlay (*LCaggregate*). The *LCaggregate* demonstrated a core of voxels that, across publications, were consistently judged to belong to the LC (see dark-blue areas in Figure 1), surrounded by a cloud of ambiguous, low-agreement voxels (see yellow areas in Figure 1). Topographically, consensus was lowest in most rostral and caudal areas (see inlay in Figure 1).

Pruning low-agreement voxels yielded a high-confidence LC volume of interest (*LCmetaMask;* available for download via: osf.io/sf2ky/) that matches the dimensions reported in post-mortem investigations (Fernandes et al., 2012) and allows assessment of the accuracy of previously published masks.

### 3.1.2. Accuracy of previously published locus coeruleus masks

Once a high-confidence LC volume of interest (*LCmetaMask*) was generated, published masks were evaluated regarding their specificity and sensitivity. That is, we assessed (1) how many voxels judged by a given mask as belonging to the locus coeruleus were part of the *LCmetaMask*, and (2) what percentage of the *LCmetaMask* was included in each individual mask. The mean across specificity and sensitivity was taken as accuracy measure to rank the masks (see Figure 2).

**Figure 2.**
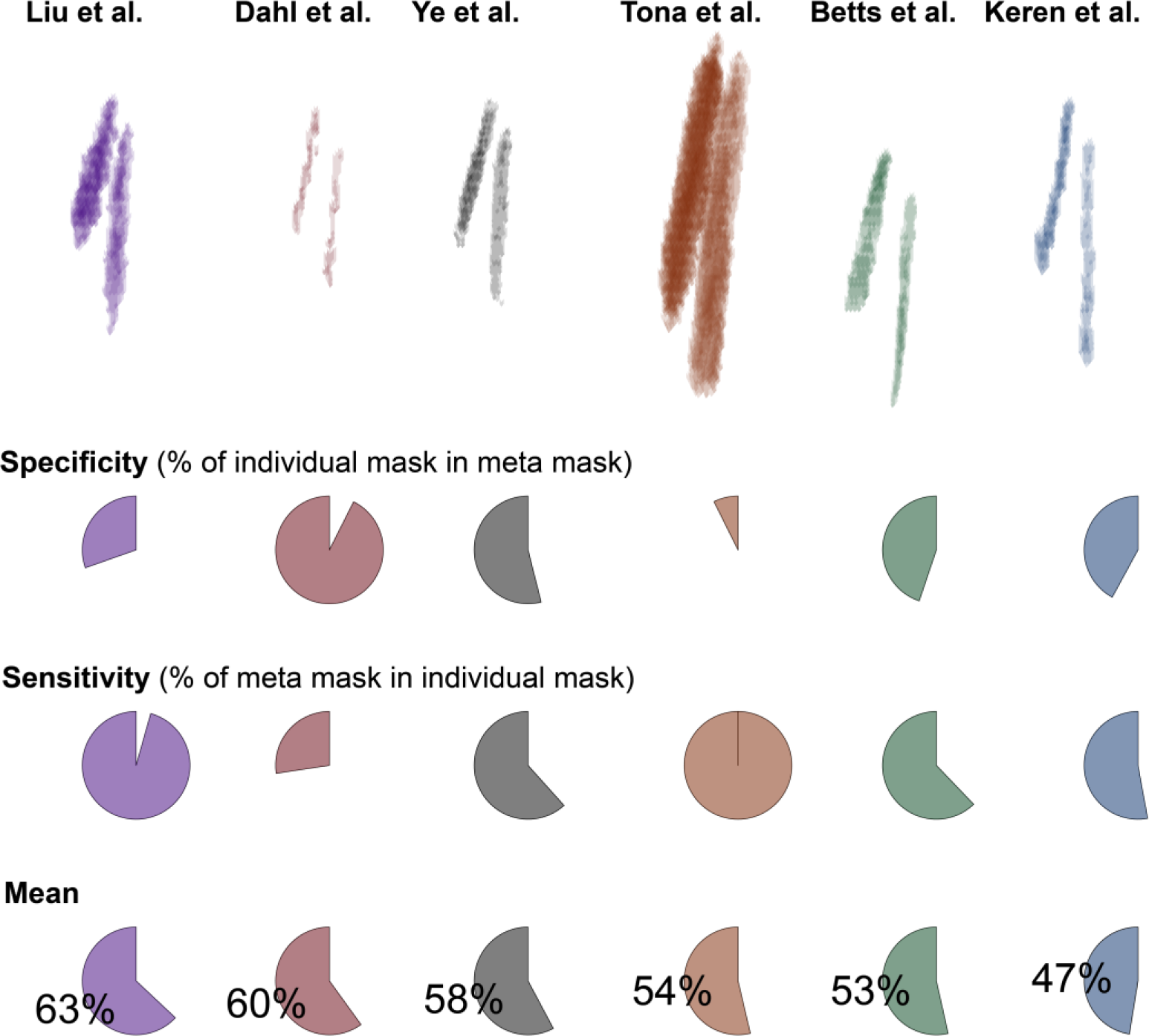
Accuracy of previously published locus coeruleus (LC) masks. Binarized masks in standard space (MNI 152, lin; top row) are ranked based on their accuracy (mean across specificity and sensitivity). Specificity expresses the percentage of a given mask that is included in the *LCmetaMask* whereas sensitivity denotes the percentage of the *LCmetaMask* that is part of each mask. For access to the individual masks, please refer to Table 1. Note that the relative position of the individual masks (top row) corresponds to their position in MNI 152 linear space; x/y/z axes are not displayed for clarity.

The masks that were computed based on the largest samples (cf. Table 1; Dahl et al., 2019; Liu et al., 2019) demonstrated the highest accuracy. Concerning specificity and sensitivity, the two masks appeared almost as mirror images. That is, the volume of interest published by Liu and colleagues evinced a high sensitivity at the cost of a relatively lower specificity (i.e., it included also non-LC voxels), while the situation was reversed for the mask by Dahl and colleagues (in line with it mapping peak LC coordinates; Dahl et al., 2019).

Notably, the two masks themselves show a high agreement (94%; Liu et al., 2019). Ye and colleagues tested a smaller sample, albeit with a higher field strength (7 T) to generate their volume of interest which appears to strike a balance between sensitivity and specificity. None of the tested masks, however, exceeded an accuracy rating of 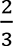, indicating that, by aggregating across publications, the *LCmetaMask* forms a volume of interest that conveys valuable information (i.e., is not redundant with previous masks).

### 3.2.1. Locus coeruleus meta mask captures high intensity voxels in independent clinical sample

We aligned and pooled across brainstem scans of participants with or known to be at-risk for ADAD mutations (cf. Dahl et al., 2019). On the group level, we observed a cluster of hyperintense voxels bordering the lateral floor of the fourth ventricle (Betts, Kirilina, et al., 2019). This LC-related hyperintensity was accurately captured by the *LCmetaMask* while excluding more medial, non-LC related high intensity voxels (see Figure 3). Taken together, this indicates that the *LCmetaMask* can be applied to independent datasets to reliably extract MR- indexed integrity across the rostrocaudal axis.

**Figure 3.**
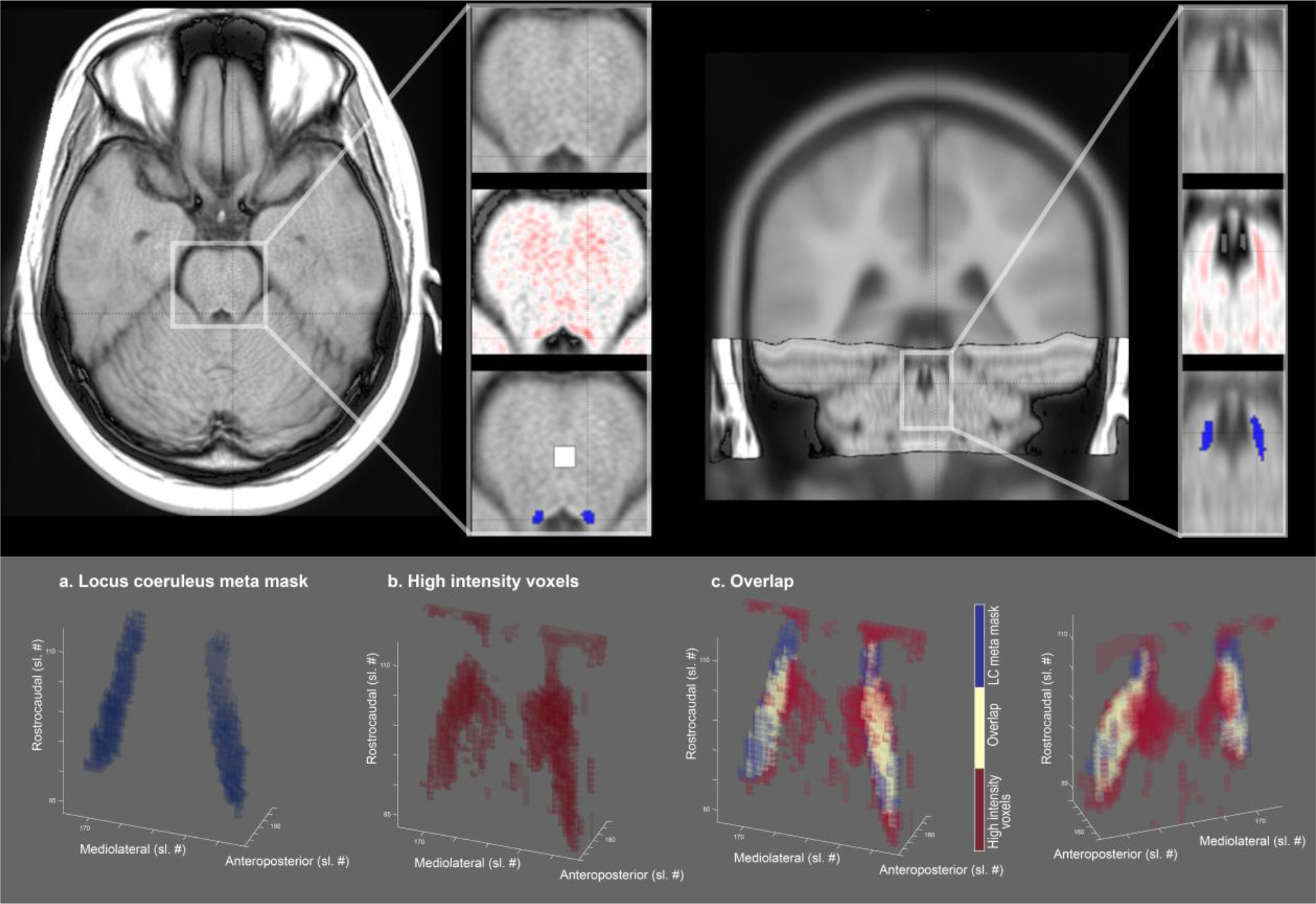
LC-related hyperintensity is accurately captured by meta mask. **Upper row**. Axial and coronal view of brainstem template overlaid on standard MNI brain. **Inlays**. The area surrounding the fourth ventricle is depicted with (1) a greyscale colormap and (2) a black-to-white-to-red colormap to highlight hyperintense areas. (3) LC-related hyperintensities closely overlap with the LC meta mask (blue overlay; white overlay, pontine reference region). **Lower row**. 3-D representation of (**a**) the LC meta mask, (**b**) above-average intensity voxels, and (**c**) their overlap from a front and back view. Sl. #, slice number in MNI 152 linear (0.5 mm) space; LC, locus coeruleus. Right hemisphere is plotted on the left.

### 3.2.2. Lower MR-indexed locus coeruleus integrity in autosomal-dominant Alzheimer’s disease

Comparing LC intensity ratios—an *in-vivo* proxy for the integrity for the structure (Keren et al., 2015)—across cognitively normal and symptomatic participants, we observed reliably lower intensity in the patient group (Wilcoxon rank sum test, *Z* = –2.177; *p* = 0.03; effect size (*r*) = –0.513; see Figure 4; mean (SD)symptomatic = 0.123 (0.051); mean (SD)cog. Normal = 0.204 (0.05)). This difference was most pronounced in middle–rostral segments of the nucleus that project to the mediotemporal lobe (Wilcoxon rank sum test of group differences in rostral vs. caudal segment; *Z* = –2.816; *p* = 0.005; effect size (*r*) = –0.532; see Figure 4). Among carriers of ADAD mutations, closer proximity to the mutation-specific median age of dementia diagnosis—termed adjusted age—was associated with lower LC ratios (mean correlation coefficient across 1,000,000 bootstraps: *rho* = –0.671; *p* = 0.024; see Figure 4).

**Figure 4.**
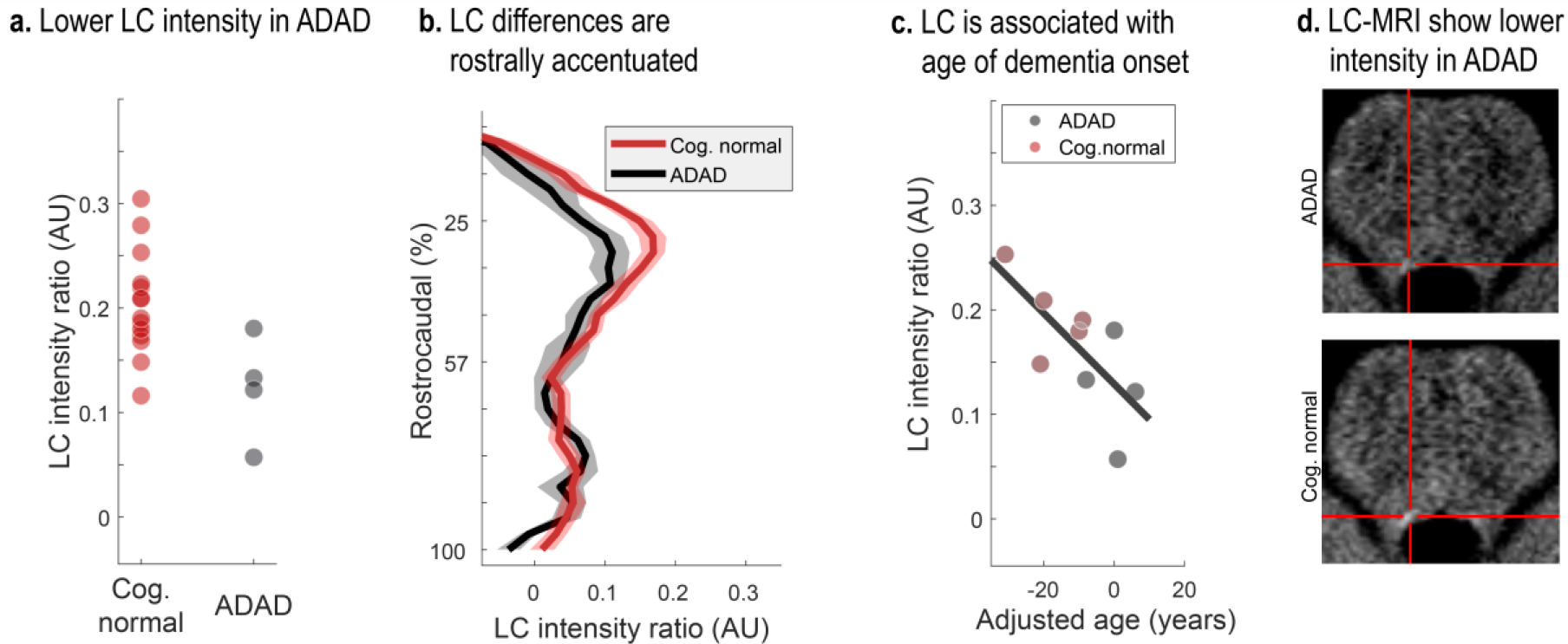
Lower LC intensity ratios in autosomal-dominant Alzheimer’s disease (ADAD). Cognitively normal (n = 14) participants are compared against mutation carriers that show cognitive impairments based on the Washington University Clinical Dementia Rating Scale (i.e., CDR > 0; n = 4). **a**. Symptomatic participants show lower overall LC intensity ratios relative to cognitively normal controls (*Z* = –2.177; *p* = 0.03). **b**. Group differences are most pronounced in middle–rostral segments of the LC (plotted on the y-axis; *Z* = –2.816; *p* = 0.005; shaded areas indicate ± 1 standard error of the mean [SEM]). **c**. Closer proximity to the mutation-specific median age of dementia diagnosis (adjusted age) is associated with lower overall LC ratios (mean correlation coefficient across 1,000,000 bootstraps: *rho* = –0.671; *p* = 0.024). Note that in panel **c** only confirmed mutation carriers (n = 9) with and without cognitive symptoms are included (black and red markers, respectively). **d**. LC-MRI of cognitively normal and symptomatic participants (four per group) were standardized and averaged. The crosshair highlights lower LC intensity in symptomatic participants. LC, locus coeruleus; Cog. normal, cognitively unimpaired participants.

Taken together, corroborating earlier post-mortem work (in LOAD; Lyness et al., 2003), we observed lower MR-indexed LC integrity in middle–rostral segments of the nucleus, potentially indicating noradrenergic neurodegeneration.

### 3.2.3. MR-indexed locus coeruleus integrity is associated with cortical tau burden

Previous animal research suggests that noradrenergic neurodegeneration may exacerbate neural decline and contribute to Alzheimer’s pathogenesis (Chalermpalanupap et al., 2018; Rorabaugh et al., 2017). Thus, we next tested whether LC intensity would be associated with cortical tau burden, a hallmark of AD.

Across cortical regions (Desikan et al., 2006), symptomatic participants demonstrated higher flortaucipir SUVR (see Figure 5). Importantly, leveraging a multivariate statistical approach (partial least squares correlation [PLSC]; Krishnan et al., 2011), we revealed a topographical pattern of LC-related tau pathology (*p* = 0.037; see Figure 5). That is, we extracted a latent variable (latent PLS score) that optimally captures the multivariate association between participants’ LC intensity and regional tau burden (*r* = 0.54 [95% confidence interval (CI): 0.167, 0.806]; lower LC intensity was associated with higher flortaucipir SUVR). Especially tau pathology in occipito-temporo-parietal regions contributed to this latent variable, as indicated by reliable bootstrap ratios (BSR; < –3; see Figure 5; BSR can be interpreted akin to Z-values; please note that we opted for a conservative threshold [– 3] due to the large number of cortical ROI). For associations of LC intensity and regional tau burden, see Table S1 in the supplementary information. In sum, in line with research in genetically modified animals (Chalermpalanupap et al., 2018; Rorabaugh et al., 2017), we observed a prominent association between in-vivo proxies of LC integrity and tau pathology.

**Figure 5.**
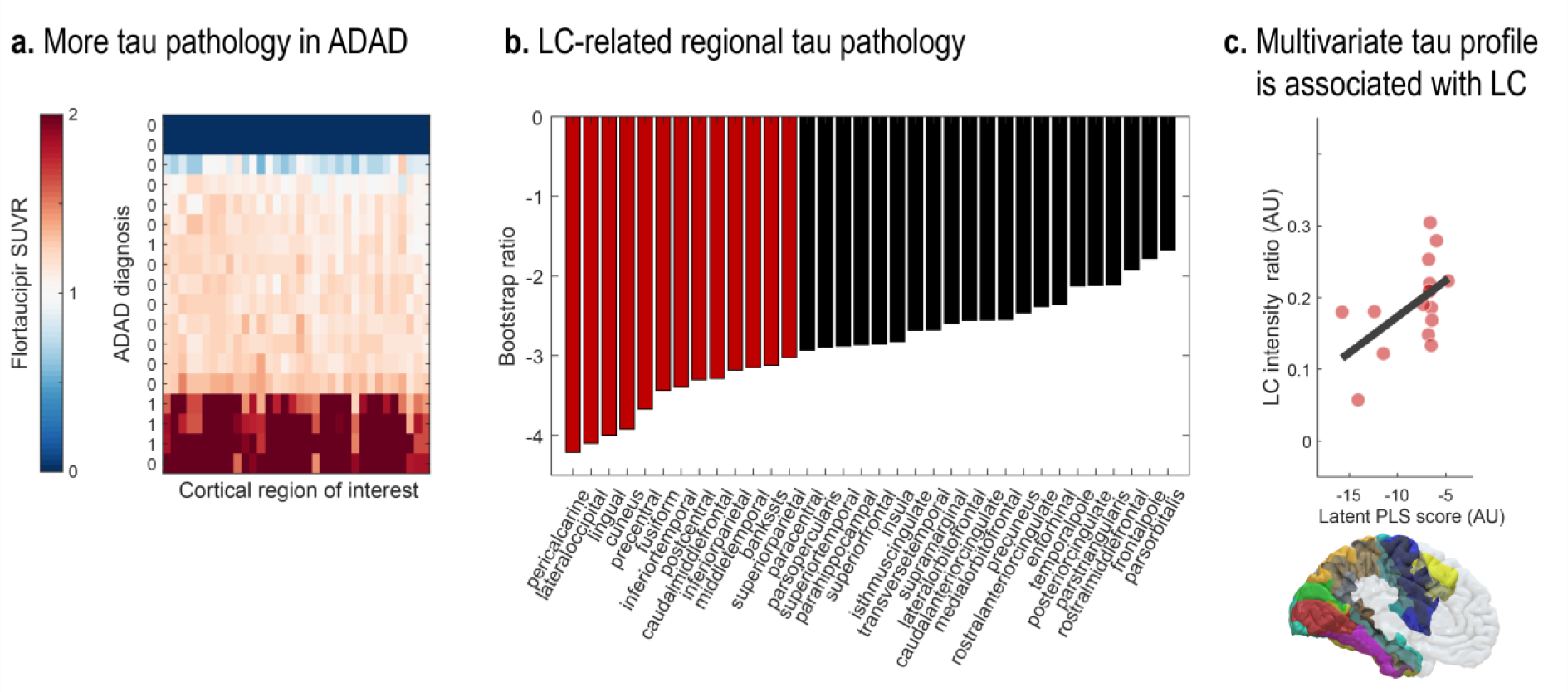
Lower LC intensity is associated with tau burden in posterior brain regions. **a**. Symptomatic participants show higher flortaucipir standardized uptake value ratios (SUVR) across most cortical regions relative to cognitively normal controls (missing PET data for two participants [dark blue bars]). **b**. Contribution of individual cortical regions of interest to a partial least squares (PLS) latent variable indexing LC-related tau pathology (Bootstrap ratios < –3 are considered reliable). **c**. Scatter plot depicting the relation between latent PLS scores and LC intensity (*p* = 0.037; *r* = 0.54 [95% CI: 0.167, 0.806]). LC, locus coeruleus; ADAD, autosomal-dominant Alzheimer’s disease.

### 3.2.4. MR-indexed locus coeruleus integrity is associated with cognitive deficits

Finally, we set out to test the behavioral relevance of our *in-vivo* proxy of LC integrity. Noradrenaline release from the LC is strongly implicated in the modulation of attentional and mnemonic processing (Berridge & Waterhouse, 2003; Dahl et al., 2020; T.-H. Lee et al., 2018; Sara, 2009). Accordingly, recent imaging studies reported positive associations between LC intensity and cognition in healthy aging (Dahl et al., 2019; Hämmerer et al., 2018; Liu et al., 2020). However, whether this relation extends to ADAD of young onset is currently unknown.

Across a range of cognitive tasks, we detected worse cognitive abilities and higher dementia symptoms in the ADAD group relative to controls (see Figure 6). Reliable group differences (Wilcoxon rank sum test; Z < –1.96 | Z > 1.96) were observed for screening instruments (the Montreal Cognitive Assessment [MoCA, Nasreddine et al., 2005]; the Cognitive Abilities Screening Instrument [CASI; Teng et al., 1994], and the Washington University Clinical Dementia Rating Scale [CDR; measure: sum of boxes; cf. O’Bryant et al., 2008]). In addition, symptomatic participants performed significantly worse on the Spanish English Verbal Learning Test (SEVLT; measures: average performance over learning trials and delayed recall; González, Mungas, & Haan, 2002), in line with reports of verbal learning tests to convey information about participants’ current and future cognitive status (Albert et al., 2001; Belleville et al., 2017; Moradi et al., 2017; Schoenberg et al., 2006).

**Figure 6.**
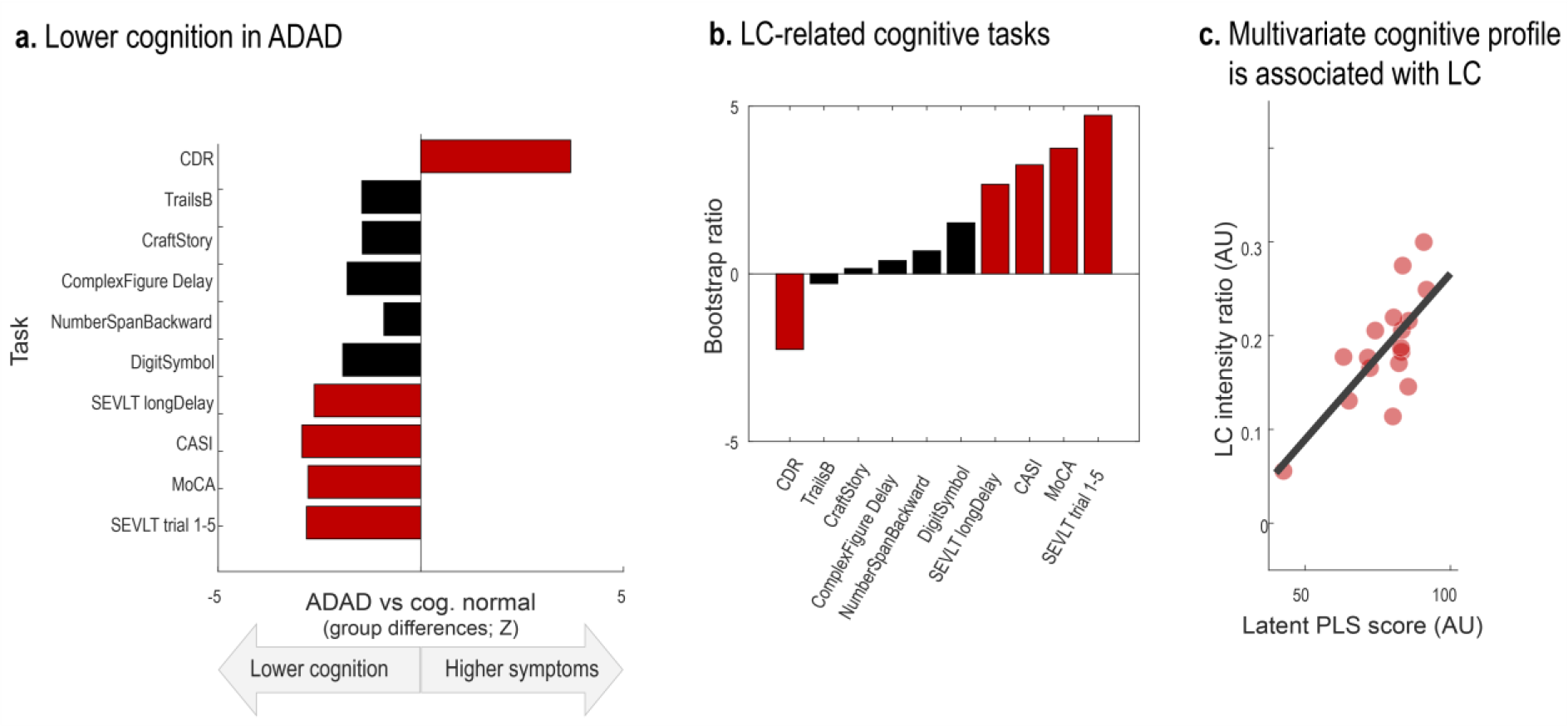
Lower LC intensity is associated with cognitive deficits. **a**. Symptomatic participants show lower cognitive performance as indicated from screening instruments for mild cognitive impairment and dementia (Clinical Dementia Rating Scale, sum of boxes [CDR]; Cognitive Abilities Screening Instrument [CASI]; Montreal Cognitive Assessment [MoCA]) as well as the Spanish English Verbal learning task (SEVLT). **b**. Contribution of individual neuropsychological tests to a partial least squares (PLS) latent variable indexing LC-related cognitive deficits (Bootstrap ratios < –1.96 and > 1.96 are considered reliable). **c**. Scatter plot depicting the relation between latent PLS scores and LC intensity (*p* = 0.016; *r* = 0.728 [95% CI: 0.408, 0.888]). LC, locus coeruleus; ADAD, autosomal-dominant Alzheimer’s disease. For references to the cognitive tasks, please refer to the main text.

We applied a partial least squares correlation (PLSC) to isolate the pattern of cognitive impairment reliably linked to MR-indexed LC integrity. Our analyses isolated a single latent variable (*p* = 0.016) that optimally expresses the multivariate association between participants’ LC intensity and cognitive performance (*r* = 0.728 [95% CI: 0.408, 0.888]). All cognitive tasks that were sensitive to distinguish symptomatic from cognitively normal participants contributed reliably to this latent variable (bootstrap ratios [BSR]< –1.96 | BSR > 1.96; see Figure 6). To conclude, across a range of neuropsychological tests, higher MR- indexed LC integrity was observed in participants with unimpaired cognitive performance.

### 3.3. Neuropathological findings in the locus coeruleus

Descriptions of the neuropathological changes occurring in the brainstem in ADAD are scarce. To characterize the pathology underlying the LC signal on MRI in ADAD, LC histopathology was compared between four carriers of the A431E mutation in *PSEN1* (aged 39–59 years, 2 ♀), including one person homozygous for the A431E mutation (Parker et al., 2019), and five neurologically normal controls (aged 37–58 years, 2 ♀). All A431E mutation carriers demonstrated severe loss of LC neurons and background astrogliosis by GFAP relative to controls. The remaining LC neurons from the A431E mutant carriers demonstrated a severe loss of neuromelanin and reactivity (see Figure 7). AT8 staining showed neuritic changes in all A431E mutation carriers with three of four showing neurofibrillary tangle formation. On staining with 4G8, three mutation carriers showed diffuse extracellular neuritic and coarse grained or compact plaques (see Figure 8). Two of five controls showed rare coarse grained extracellular plaques. Cotton-wool amyloid plaques are large ball-like extracellular plaques that displace nearby structures and are commonly seen in autosomal dominant forms of Alzheimer’s disease. The A431E homozygote case has a higher density of these plaques throughout the brain and brainstem relative to the heterozygote cases. Alpha synuclein staining did not reveal any Lewy body pathology in any samples. Immunostaining using antibodies against 4-repeat tau (RD4) and 3-repeat-tau (RD3) showed positivity for both isoforms.

**Figure 7.**
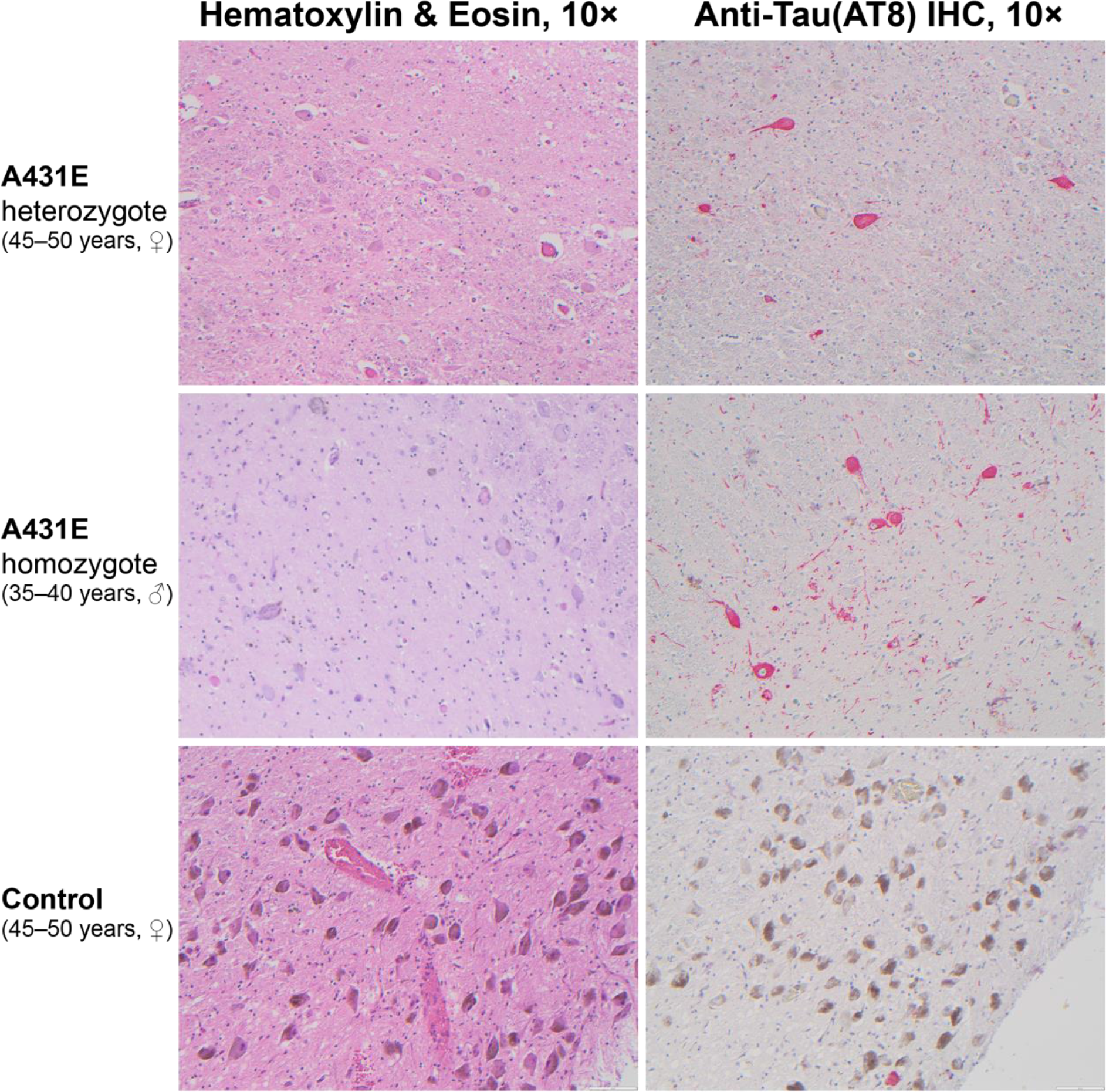
Neuropathological findings in the locus coeruleus of a 45–50 year old woman heterozygous for the A431E mutation in *PSEN1*, a 35–40 year old man homozygous for the A431E mutation in PSEN1, and a normal control (45–50 year old). Hematoxylin and Eosin (H&E) staining shows profound drop-out of neurons in the *PSEN1* mutation carriers compared to control. Neurons within carriers lack neuromelanin and show loss of pigment; whereas, neurons within control show robust pigmentation and neuromelanin expression. Immunostained slides with anti-tau (AT8) antibodies using red chromagen demonstrate neurofibrillary tangles within neurons and the presence of many tau positive threads in *PSEN1* mutation carriers with only a very rare single neurofibrillary tangle observed in normal control (lower portion of image).

**Figure 8.**
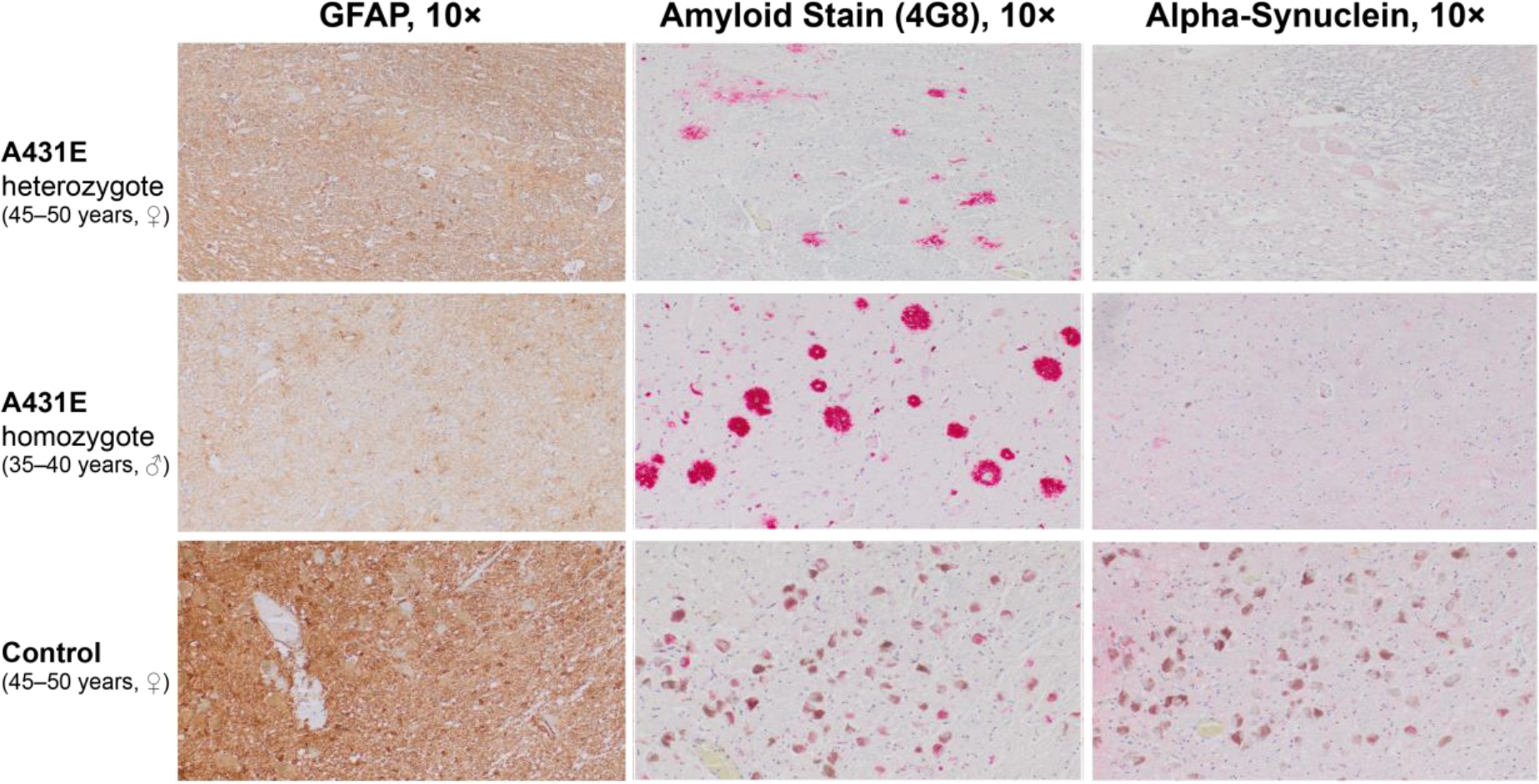
Neuropathological findings in the locus coeruleus of a 45–50 year old woman heterozygous for the A431E mutation in PSEN1, a 35–40 year old man homozygous for the A431E mutation, and a normal control (45–50 years old). GFAP highlights the background reactive astrogliosis surrounding the locus coeruleus with the homozygous case demonstrating the most reactive background and increased number of astrocytes with cell body and process hypertrophy. The amyloid stain (4G8) highlights the extracellular amyloid plaques in the locus coeruleus with the heterozygote case demonstrating diffuse and compact amyloid plaques. The homozygote case demonstrates large cotton-wool amyloid plaques. The control does not have any amyloid plaques in the area. The alpha-synuclein did not reveal Lewy body pathology.

## 4. Discussion

Animal and post-mortem research indicates that the brain’s noradrenergic system plays a central role in the pathogenesis of Alzheimer’s disease (Braak et al., 2011; Weinshenker, 2018). *In vivo* investigations, however, have long been hindered by challenges in reliable non-invasive LC assessments (Astafiev et al., 2010; Keren et al., 2009). While initial intriguing *in vivo* studies in late-onset Alzheimer’s are underway (LOAD; e.g., Betts, Cardenas-Blanco, et al., 2019; Jessen et al., 2018), unequivocally distinguishing age- from disease-related LC alterations is difficult on this basis (Dahl et al., 2019; Liu et al., 2019).

Focusing instead on types of dementia developing earlier in life provides a unique window into the mechanisms driving Alzheimer’s development (ADAD; Bateman et al., 2012; Jacobs, Becker, et al., 2019; Ringman, 2005).

Leveraging a meta-analytical approach, we first synthesized LC localizations and dimensions across previously published studies to improve the reliability and validity of MR-based LC detection. In a second step, we applied this newly generated volume of interest to determine whether MR-indexed LC integrity can serve as marker for noradrenergic degeneration in ADAD and verified this pathology using post-mortem samples.

Our analyses isolated a cluster of high-confidence voxels that (1) is consistently identified as LC across published studies and (2) corresponds well to post-mortem reports (Fernandes et al., 2012). Next, exploiting this refined spatial characterization we semi-automatically extracted intensity data in an independent sample of participants with or known to be at-risk for mutations in genes associated with ADAD. We revealed lower MR-indexed LC integrity in symptomatic mutation carriers. Group differences were most pronounced in middle to rostral segments of the LC (Ehrenberg et al., 2017; Lyness et al., 2003) and scaled with proximity to the mutation-specific median age of dementia diagnosis (Ryman et al., 2014). Beyond that, lower overall LC intensity was linked to higher tau burden in posterior brain regions (Chalermpalanupap et al., 2018) and worse cognitive performance across several neuropsychological tests (Dahl et al., 2019; Liu et al., 2020). Taken together, our finding of diminished LC integrity in ADAD suggests a prominent role of the noradrenergic system in this neurodegenerative disease.

### 4.1. Aggregating across published locus coeruleus maps yields high confidence meta mask

Imaging of small brainstem structures that are not visible using conventional MR sequences is notoriously challenging (for discussions, see Astafiev et al., 2010; Keren et al., 2009) and warrants a close alignment of individual scans (for recommendations, see Betts, Kirilina, et al., 2019; Liu et al., 2017). Here we used Advanced Normalization Tools (ANTs; Avants et al., 2011, 2009) for brainstem normalization. ANTs is becoming increasingly recognized as a powerful tool for accurate coregistrations – it has won several internationally recognized medical image processing challenges (e.g., Klein et al., 2009 for a full list see github.com/ANTsX/ANTs) and was used in all of the currently best ranked LC mapping studies (3/3, [relative to 1/3 of the remaining studies]; Dahl et al., 2019; Liu et al., 2019; Ye et al., 2021).

Besides normalization, LC mapping studies employing larger sample sizes and a higher MR-field strength evinced a higher accuracy, as indicated by spatial congruence with the meta mask (Dahl et al., 2019; Liu et al., 2019; Ye et al., 2021). We hope that by sharing an unbiased, biologically-plausible LC volume of interest that is based on more than 1,000 participants and all commonly used sequence types (FSE, MT, FLASH; see Table 1) we assist future functional and structural imaging studies in classifying brainstem effects (this LC meta mask is available for download via osf.io/sf2ky/).

Moreover, the LC meta mask can be combined with a template-based semi-automatic LC intensity assessment to systematically compare LC integrity in patients and controls (for a step-by-step description of the procedure, see (Dahl et al., 2019); for an application, see e.g., (Gallant et al., 2021)). In short, LC-MRI are standardized to MNI space and masked with the LC volume of interest. After masking, the relative intensity within the LC (i.e., standardized with respect to a reference area) can be compared across groups.

#### 4.2.1. Lower MR-indexed locus coeruleus integrity in autosomal-dominant Alzheimer’s disease

Applying the newly generated LC meta mask to an independent clinical sample, we demonstrate its utility for accurately extracting intensity information across the rostrocaudal extent of the nucleus. We observed lower middle–rostral LC intensity in symptomatic carriers of ADAD-causing mutations. This finding is consistent with post-mortem reports of substantial noradrenergic neurodegeneration in LOAD (Lyness et al., 2003). Some previous post-mortem studies in ADAD also noted disproportionate depigmentation and neuronal loss in the LC (Janssen et al., 2000; Snider et al., 2005) which we verified in four persons dying with the A431E mutation in *PSEN1*. *In vivo* imaging studies are in general agreement with the histological findings and indicate lower LC contrast in LOAD patients (Betts, Cardenas-Blanco, et al., 2019; Dordevic et al., 2017; Takahashi et al., 2015). To date there is only a single imaging study focusing on early-onset Alzheimer’s (ADAD; Jacobs, Becker, et al., 2019), albeit in non-symptomatic mutation carriers (i.e., at a preclinical stage). Our observation that LC intensity scaled with proximity of dementia onset (adjusted age) across non-symptomatic and symptomatic mutation carriers matches a negative age association in mutation carriers reported by Jacobs and colleagues (2019) and might indicate a progressive deterioration. In conjunction with prior post-mortem validations of LC MRI (Keren et al., 2015; Cassidy et al., 2019) and decreased cell counts in LOAD (Lyness et al., 2003), our post-mortem validation of decreased cell counts in the LC in ADAD are indicative of noradrenergic neurodegeneration in ADAD.

#### 4.2.2. MR-indexed locus coeruleus integrity is associated with cortical tau burden

Leveraging a multivariate statistical approach, we further revealed a pattern of LC-related tau pathology that was most pronounced in posterior brain regions (cf. Jacobs, Becker, et al., 2019). This observation supports the notion that loss of noradrenergic neurons may exacerbate the severity of other Alzheimer’s hallmarks, like tau pathology (Chalermpalanupap et al., 2018). The LC is one of the first brain regions in which aberrant tau can be detected at relatively early ages (i.e., the first decades of life; Braak et al., 2011). The accumulation of abnormal tau has been hypothesized to shift noradrenergic cells to a mode of hyperactivity which in turn promotes tau release and spread along LC’ widespread axonal pathways (Chalermpalanupap et al., 2017; Weinshenker, 2018). While tau gradually propagates through cortical regions (categorized as Braak stages), noradrenergic neurons first shrink in size and then degenerate (Ehrenberg et al., 2017; Kelly et al., 2017; Theofilas et al., 2017). The frank decline of noradrenergic cells is preceded by a period of dysfunctional neurotransmission in LC’s terminal regions like the mediotemporal lobe (Weinshenker, 2018). Thus, the negative association between *in vivo* proxies for LC integrity and tau burden may signify a more advanced Alzheimer’s disease stage including noradrenergic neurodegeneration and wide- spread tau pathology.

#### 4.2.3. MR-indexed locus coeruleus integrity is associated with cognitive deficits

Finally, we found evidence for an association between MR-indexed LC integrity and cognitive performance across several neuropsychological tests. Noradrenergic neuromodulation has been implicated in a range of cognitive functions, prominently including attention and memory (Berridge & Waterhouse, 2003; Bouret & Sara, 2005; Corbetta et al., 2008; Mather et al., 2016; Sara, 2009). In particular, mediated via β-adrenoceptors, noradrenaline release promotes long-term-potentiation in the hippocampus—a key determinant of synaptic plasticity and memory (O’Dell et al., 2015). Recent optogenetic research moreover indicates a causal role of LC activity in memory acquisition and consolidation, potentially supported by co-release of dopamine (Duszkiewicz et al., 2019; Takeuchi et al., 2016; Uematsu et al., 2017; Wagatsuma et al., 2018). Consistent with these observations in animals, recent *in vivo* human research demonstrated an association between MR-indexed LC integrity and memory performance in healthy aging (Dahl et al., 2019; Elman et al., 2021; Hämmerer et al., 2018; Liu et al., 2020). Extending evidence from healthy populations, we here confirm the behavioral relevance of MR-indexed LC integrity in tracking cognitive decline in a clinical sample. Neuropsychological tests that proved sensitive to distinguish patients from controls were reliably linked to LC integrity (González et al., 2002; Nasreddine et al., 2005; O’Bryant et al., 2008; Teng et al., 1994). In line with reports indicating verbal learning tests as effective early markers of Alzheimer’s pathology (Albert et al., 2001; Belleville et al., 2017; Moradi et al., 2017; Schoenberg et al., 2006), verbal learning and memory showed the closest link to LC integrity (cf. Dahl et al., 2019, for analogous results in later life). Taken together, our findings suggest that noradrenergic neurotransmission supports memory performance whereas dysfunctions therein are associated with Alzheimer’s related cognitive decline.

### 4.3. Limitations and conclusions

Some limitations should be noted. First, the contrast mechanisms underlying LC-MRI constitute an active area of research (for a discussion, see Betts, Kirilina, et al., 2019) and future studies may help determine what precise physiological processes are reflected in MR-contrast differences. Second, as a cross-sectional observational study, it is not possible to conclude a causative relationship between loss of LC integrity and cognitive symptoms and deposition of abnormal tau in persons with ADAD. Finally, with a prevalence of about 5 cases per 100,000 persons at risk, ADAD is a rare disease (Campion et al., 1999) accounting for less than 1% of all Alzheimer’s cases (Bateman et al., 2011). All but one ADAD mutation carriers in the current study had the same mutation in *PSEN1* (A431E) which has specific features (e.g., co-occurring spastic paraparesis and possibly atypical posterior-predominant tau deposition) that may limit generalizability of the current findings. While providing insights into the pathogenesis of Alzheimer’s largely disentangled from factors associated with aging, the presented findings need to be corroborated by longitudinal evidence in larger samples (Lindenberger et al., 2011).

In conclusion, we applied a meta-analytical approach to advance the reliability and validity of MR-based LC detection. Using non-invasive *in vivo* proxies, we revealed lower LC integrity in symptomatic carriers of ADAD-causing mutations. Moreover, LC integrity was strongly associated with cortical tau pathology and memory decline. In summary, this provides support for a prominent role of the noradrenergic system in Alzheimer’s disease (Mather & Harley, 2016; Weinshenker, 2018).

## Data Availability

The here described LC meta mask as well as pontine reference mask is available for download on an Open Science Framework repository (osf.io/sf2ky/). All sources for previously published masks are listed in Table 1. For access to raw imaging and clinical data, please refer to the HCP (http://nda.nih.gov/ccf).

https://nda.nih.gov

## Acknowledgements

The authors thank all study participants for their contribution.

## 7 Supplementary information

**Table S1:** Associations of locus coeruleus intensity and regional tau burden

| Region of interest<sup>1</sup> | *rho* | *p* |
| --- | --- | --- |
| bankssts | -0.688 | 0.004 |
| transversetemporal | -0.656 | 0.007 |
| postcentral | -0.641 | 0.009 |
| inferiorparietal | -0.641 | 0.009 |
| inferiortemporal | -0.638 | 0.009 |
| cuneus | -0.624 | 0.012 |
| fusiform | -0.609 | 0.014 |
| parstriangularis | -0.582 | 0.02 |
| pericalcarine | -0.553 | 0.029 |
| posteriorcingulate | -0.544 | 0.032 |
| precuneus | -0.532 | 0.036 |
| caudalmiddlefrontal | -0.526 | 0.038 |
| entorhinal | -0.524 | 0.04 |
| lingual | -0.509 | 0.046 |
| middletemporal | -0.509 | 0.046 |
| isthmuscingulate | -0.503 | 0.049 |
| temporalpole | -0.497 | 0.052 |
| lateraloccipital | -0.494 | 0.054 |
| caudalanteriorcingulate | -0.488 | 0.057 |
| rostralanteriorcingulate | -0.476 | 0.064 |
| superiorparietal | -0.459 | 0.076 |
| paracentral | -0.459 | 0.076 |
| parahippocampal | -0.456 | 0.078 |
| insula | -0.441 | 0.089 |
| lateralorbitofrontal | -0.435 | 0.094 |
| parsorbitalis | -0.418 | 0.109 |
| frontalpole | -0.409 | 0.117 |
| parsopercularis | -0.403 | 0.123 |
| precentral | -0.382 | 0.145 |
| medialorbitofrontal | -0.376 | 0.151 |
| superiortemporal | -0.365 | 0.165 |
| superiorfrontal | -0.312 | 0.239 |
| rostralmiddlefrontal | -0.282 | 0.288 |
| supramarginal | -0.232 | 0.385 |

*Note:* <sup>1</sup>Parcellation based on Desikan et al., (2006).
Tau data was averaged across hemispheres before analysis (see *Material and methods*).
*Rho* and *p* values are based on non-parametric Spearman's correlations (*n* = 16).

**Table S2:** Associations of locus coeruleus intensity and cognitive performance

| Cognitive task | *rho* | *p* |
| --- | --- | --- |
| Craft Story | 0.62 | 0.008 |
| Complex Figure Delay | 0.589 | 0.013 |
| Trails B | 0.498 | 0.042 |
| Number Span Backward | 0.459 | 0.064 |
| SEVLT_total (trial 1–5) | 0.447 | 0.072 |
| CASI | 0.286 | 0.265 |
| SEVLT long delay | 0.006 | 0.981 |
| MoCA | -0.02 | 0.94 |
| Digit Symbol | -0.16 | 0.539 |
| CDR | -0.405 | 0.107 |

*Note: Rho and p values are based on non-parametric Spearman's correlations (n = 17). See* *Material and methods* for a description of the cognitive tasks.

## References

1. Albert, M. S., Moss, M. B., Tanzi, R., & Jones, K. (2001). Preclinical prediction of AD using neuropsychological tests. Journal of the International Neuropsychological Society, 7(5), 631–639. https://doi.org/10.1017/S1355617701755105

2. Alzheimer’s Disease International. (2019). World Alzheimer Report 2019: Attitudes to Dementia. Alzheimer’s Disease International: London.

3. Arendt, T., Brückner, M. K., Morawski, M., Jäger, C., & Gertz, H. J. (2015). Early neurone loss in Alzheimer’s disease: cortical or subcortical? Acta Neuropathologica Communications, 3, 10. https://doi.org/10.1186/s40478-015-0187-1

4. Astafiev, S. V., Snyder, A. Z., Shulman, G. L., & Corbetta, M. (2010). Comment on “Modafinil shifts human locus coeruleus to low-tonic, high-phasic activity during functional MRI” and “Homeostatic sleep pressure and responses to sustained attention in the suprachiasmatic area.” Science, 328(5976), 309. https://doi.org/10.1126/science.1177200

5. Avants, B. B., Tustison, N. J., Song, G., Cook, P. A., Klein, A., & Gee, J. C. (2011). A reproducible evaluation of ANTs similarity metric performance in brain image registration. NeuroImage, 54(3), 2033–2044. https://doi.org/10.1016/j.neuroimage.2010.09.025

6. Avants, B. B., Tustison, N., & Song, G. (2009). Advanced Normalization Tools: V1.0. Insight Journal, 2. http://hdl.handle.net/10380/3113

7. Bateman, R. J., Aisen, P. S., De Strooper, B., Fox, N. C., Lemere, C. A., Ringman, J. M., Salloway, S., Sperling, R. A., Windisch, M., & Xiong, C. (2011). Autosomal-dominant Alzheimer’s disease: A review and proposal for the prevention of Alzheimer’s disease. In Alzheimer’s Research and Therapy (Vol. 2, Issue 6, p. 1). BioMed Central. https://doi.org/10.1186/alzrt59

8. Bateman, R. J., Xiong, C., Benzinger, T. L. S., Fagan, A. M., Goate, A., Fox, N. C., Marcus, D. S., Cairns, N. J., Xie, X., Blazey, T. M., Holtzman, D. M., Santacruz, A., Buckles, V., Oliver, A., Moulder, K., Aisen, P. S., Ghetti, B., Klunk, W. E., McDade, E., … Morris, J. C. (2012). Clinical and biomarker changes in dominantly inherited Alzheimer’s disease. New England Journal of Medicine, 367(9), 795–804. https://doi.org/10.1056/NEJMoa1202753

9. Belleville, S., Fouquet, C., Hudon, C., Zomahoun, H. T. V., & Croteau, J. (2017). Neuropsychological measures that predict progression from mild cognitive impairment to Alzheimer’s type dementia in older adults: A systematic review and meta-analysis. Neuropsychology Review, 27(4), 328–353. https://doi.org/10.1007/s11065-017-9361-5

10. Berridge, C. W., & Waterhouse, B. D. (2003). The locus coeruleus-noradrenergic system: Modulation of behavioral state and state-dependent cognitive processes. Brain Research Reviews, 42(1), 33–84. https://doi.org/10.1016/S0165-0173(03)00143-7

11. Bertram, L., Lill, C. M., & Tanzi, R. E. (2010). The genetics of alzheimer disease: Back to the future. In Neuron (Vol. 68, Issue 2, pp. 270–281). Neuron. https://doi.org/10.1016/j.neuron.2010.10.013

12. Betts, M. J., Cardenas-Blanco, A., Kanowski, M., Jessen, F., & Düzel, E. (2017). In vivo MRI assessment of the human locus coeruleus along its rostrocaudal extent in young and older adults. NeuroImage, 163, 150–159. https://doi.org/10.1016/j.neuroimage.2017.09.042

13. Betts, M. J., Cardenas-Blanco, A., Kanowski, M., Spottke, A., Teipel, S. J., Kilimann, I., Jessen, F., & Düzel, E. (2019). Locus coeruleus MRI contrast is reduced in Alzheimer’s disease dementia and correlates with CSF Aβ levels. *Alzheimer’s & Dementia: Diagnosis*, Assessment & Disease Monitoring, 11, 281–285. https://doi.org/10.1016/j.dadm.2019.02.001

14. Betts, M. J., Kirilina, E., Otaduy, M. C. G., Ivanov, D., Acosta-Cabronero, J., Callaghan, M. F., Lambert, C., Cardenas-Blanco, A., Pine, K., Passamonti, L., Loane, C., Keuken, M. C., Trujillo, P., Lüsebrink, F., Mattern, H., Liu, K. Y., Priovoulos, N., Fliessbach, K., Dahl, M. J., … Hämmerer, D. (2019). Locus coeruleus imaging as a biomarker for noradrenergic dysfunction in neurodegenerative diseases. Brain, 142(9), 2558–2571. https://doi.org/10.1093/brain/awz193

15. Bouret, S., & Sara, S. J. (2005). Network reset: A simplified overarching theory of locus coeruleus noradrenaline function. Trends in Neurosciences, 28(11), 574–582. https://doi.org/10.1016/j.tins.2005.09.002

16. Braak, H., & Del Tredici, K. (2016). Potential Pathways of Abnormal Tau and α-Synuclein Dissemination in Sporadic Alzheimer’s and Parkinson’s Diseases. Cold Spring Harbor Perspectives in Biology, 8(11), a023630. https://doi.org/10.1101/CSHPERSPECT.A023630

17. Braak, H., Thal, D. R., Ghebremedhin, E., & Del Tredici, K. (2011). Stages of the pathologic process in Alzheimer disease: Age categories from 1 to 100 years. Journal of Neuropathology and Experimental Neurology, 70(11), 960–969. https://doi.org/10.1097/NEN.0b013e318232a379

18. Brookmeyer, R., Johnson, E., Ziegler-Graham, K., & Arrighi, H. M. (2007). Forecasting the global burden of Alzheimer’s disease. Alzheimer’s & Dementia, 3(3), 186–191. https://doi.org/10.1016/J.JALZ.2007.04.381

19. Campion, D., Dumanchin, C., Hannequin, D., Dubois, B., Belliard, S., Puel, M., Thomas- Anterion, C., Michon, A., Martin, C., Charbonnier, F., Raux, G., Camuzat, A., Penet, C., Mesnage, V., Martinez, M., Clerget-Darpoux, F., Brice, A., & Frebourg, T. (1999). Early- onset autosomal dominant Alzheimer disease: Prevalence, genetic heterogeneity, and mutation spectrum. American Journal of Human Genetics, 65(3), 664–670. https://doi.org/10.1086/302553

20. Canter, R. G., Penney, J., & Tsai, L. H. (2016). The road to restoring neural circuits for the treatment of Alzheimer’s disease. In Nature (Vol. 539, Issue 7628, pp. 187–196). Nature Publishing Group. https://doi.org/10.1038/nature20412

21. Cassidy, C. M., Zucca, F. A., Girgis, R. R., Baker, S. C., Weinstein, J. J., Sharp, M. E., Bellei, C., Valmadre, A., Vanegas, N., Kegeles, L. S., Brucato, G., Jung Kang, U., Sulzer, D., Zecca, L., Abi-Dargham, A., & Horga, G. (2019). Neuromelanin-sensitive MRI as a noninvasive proxy measure of dopamine function in the human brain. Proceedings of the National Academy of Sciences of the United States of America, 116(11), 5108–5117. https://doi.org/10.1073/pnas.1807983116

22. Chalermpalanupap, T., Schroeder, J. P., Rorabaugh, J. M., Liles, L. C., Lah, J. J., Levey, A. I., & Weinshenker, D. (2018). Locus coeruleus ablation exacerbates cognitive deficits, neuropathology, and lethality in P301S tau transgenic mice. The Journal of Neuroscience : The Official Journal of the Society for Neuroscience, 38(1), 74–92. https://doi.org/10.1523/JNEUROSCI.1483-17.2017

23. Chalermpalanupap, T., Weinshenker, D., & Rorabaugh, J. M. (2017). Down but not out: The consequences of pretangle tau in the locus coeruleus. Neural Plasticity, 2017, 7829507. https://doi.org/10.1155/2017/7829507

24. Corbetta, M., Patel, G., & Shulman, G. L. (2008). The reorienting system of the human brain: From environment to theory of mind. Neuron, 58(3), 306–324. https://doi.org/10.1016/j.neuron.2008.04.017

25. Craft, S., Newcomer, J., Kanne, S., Dagogo-Jack, S., Cryer, P., Sheline, Y., Luby, J., Dagogo- Jack, A., & Alderson, A. (1996). Memory improvement following induced hyperinsulinemia in Alzheimer’s disease. Neurobiology of Aging, 17(1), 123–130. https://doi.org/10.1016/0197-4580(95)02002-0

26. Crary, J. F., Trojanowski, J. Q., Schneider, J. A., Abisambra, J. F., Abner, E. L., Alafuzoff, I., Arnold, S. E., Attems, J., Beach, T. G., Bigio, E. H., Cairns, N. J., Dickson, D. W., Gearing, M., Grinberg, L. T., Hof, P. R., Hyman, B. T., Jellinger, K., Jicha, G. A., Kovacs, G. G., … Nelson, P. T. (2014). Primary age-related tauopathy (PART): a common pathology associated with human aging. Acta Neuropathologica, 128(6), 755–766. https://doi.org/10.1007/s00401-014-1349-0

27. Dahl, M. J., Mather, M., Düzel, S., Bodammer, N. C., Lindenberger, U., Kühn, S., & Werkle- Bergner, M. (2019). Rostral locus coeruleus integrity is associated with better memory performance in older adults. Nature Human Behaviour, 3(11), 1203–1214. https://doi.org/10.1038/s41562-019-0715-2

28. Dahl, M. J., Mather, M., Sander, M. C., & Werkle-Bergner, M. (2020). Noradrenergic responsiveness supports selective attention across the adult lifespan. Journal of Neuroscience, 40(22), 4372–4390. https://doi.org/10.1523/JNEUROSCI.0398-19.2020

29. Dale, A. M., Fischl, B., & Sereno, M. I. (1999). Cortical surface-based analysis: I. Segmentation and surface reconstruction. NeuroImage, 9(2), 179–194. https://doi.org/10.1006/nimg.1998.0395

30. Desikan, R. S., Ségonne, F., Fischl, B., Quinn, B. T., Dickerson, B. C., Blacker, D., Buckner, R. L., Dale, A. M., Maguire, R. P., Hyman, B. T., Albert, M. S., & Killiany, R. J. (2006). An automated labeling system for subdividing the human cerebral cortex on MRI scans into gyral based regions of interest. NeuroImage, 31(3), 968–980. https://doi.org/10.1016/j.neuroimage.2006.01.021

31. Dordevic, M., Müller-Fotti, A., Müller, P., Schmicker, M., Kaufmann, J., & Müller, N. G. (2017). Optimal Cut-Off Value for Locus Coeruleus-to-Pons Intensity Ratio as Clinical Biomarker for Alzheimer’s Disease: A Pilot Study. Journal of Alzheimer’s Disease Reports, 1(1), 159–167. https://doi.org/10.3233/adr-170021

32. Duszkiewicz, A. J., McNamara, C. G., Takeuchi, T., & Genzel, L. (2019). Novelty and Dopaminergic Modulation of Memory Persistence: A Tale of Two Systems. Trends in Neurosciences, 42(2), 102–114. https://doi.org/10.1016/J.TINS.2018.10.002

33. Ehrenberg, A. J., Nguy, A. K., Theofilas, P., Dunlop, S., Suemoto, C. K., Di Lorenzo Alho, A. T., Leite, R. P., Diehl Rodriguez, R., Mejia, M. B., Rüb, U., Farfel, J. M., de Lucena Ferretti-Rebustini, R. E., Nascimento, C. F., Nitrini, R., Pasquallucci, C. A., Jacob-Filho, W., Miller, B., Seeley, W. W., Heinsen, H., & Grinberg, L. T. (2017). Quantifying the accretion of hyperphosphorylated tau in the locus coeruleus and dorsal raphe nucleus: The pathological building blocks of early Alzheimer’s disease. Neuropathology and Applied Neurobiology, 43(5), 393–408. https://doi.org/10.1111/nan.12387

34. Elman, J. A., Puckett, O. K., Beck, A., Panizzon, M. S., Sanderson-Cimino, M. E., Gustavson, D. E., Lyons, M. J., Franz, C. E., & Kremen, W. S. (2021). MRI-assessed locus coeruleus integrity is heritable and associated with cognition, Alzheimer’s risk, and sleep-wake disturbance. Alzheimer’s & Dementia, 17(6), 1017– 1025. https://doi.org/10.1002/alz.044862

35. Fernandes, P., Regala, J., Correia, F., & Gonçalves-Ferreira, A. J. (2012). The human locus coeruleus 3-D stereotactic anatomy. Surgical and Radiologic Anatomy, 34(10), 879–885. https://doi.org/10.1007/s00276-012-0979-y

36. Gallant, S. N., Kennedy, B. L., Bachman, S. L., Huang, R., Lee, T.-H., & Mather, M. (2021). Behavioral and fMRI evidence that arousal enhances bottom-up attention and memory selectivity in young but not older adults. BioRxiv, 2021.07.02.450802. https://doi.org/10.1101/2021.07.02.450802

37. Ghosh, A., Torraville, S. E., Mukherjee, B., Walling, S. G., Martin, G. M., Harley, C. W., & Yuan, Q. (2019). An experimental model of Braak’s pretangle proposal for the origin of Alzheimer’s disease: The role of locus coeruleus in early symptom development. Alzheimer’s Research and Therapy, 11(1), 59. https://doi.org/10.1186/s13195-019-0511-2

38. Goate, A., Chartier-Harlin, M. C., Mullan, M., Brown, J., Crawford, F., Fidani, L., Giuffra, L., Haynes, A., Irving, N., James, L., Mant, R., Newton, P., Rooke, K., Roques, P., Talbot, C., Pericak-Vance, M., Roses, A., Williamson, R., Rossor, M., … Hardy, J. (1991). Segregation of a missense mutation in the amyloid precursor protein gene with familial Alzheimer’s disease. Nature, 349(6311), 704–706. https://doi.org/10.1038/349704a0

39. González, H. M., Mungas, D., & Haan, M. N. (2002). A verbal learning and memory test for English- and Spanish-speaking older Mexican-American adults. Clinical Neuropsychologist, 16(4), 439–451. https://doi.org/10.1076/clin.16.4.439.13908

40. Greve, D. N., Salat, D. H., Bowen, S. L., Izquierdo-Garcia, D., Schultz, A. P., Catana, C., Becker, J. A., Svarer, C., Knudsen, G. M., Sperling, R. A., & Johnson, K. A. (2016). Different partial volume correction methods lead to different conclusions: An 18F-FDG- PET study of aging. NeuroImage, 132, 334–343. https://doi.org/10.1016/j.neuroimage.2016.02.042

41. Greve, D. N., Svarer, C., Fisher, P. M., Feng, L., Hansen, A. E., Baare, W., Rosen, B., Fischl, B., & Knudsen, G. M. (2014). Cortical surface-based analysis reduces bias and variance in kinetic modeling of brain PET data. NeuroImage, 92, 225–236. https://doi.org/10.1016/j.neuroimage.2013.12.021

42. Grinberg, L. T., & Heinsen, H. (2017). Light at the beginning of the tunnel? Investigating early mechanistic changes in Alzheimer’s disease. Brain, 140(11), 2770–2773. https://doi.org/10.1093/brain/awx261

43. Hämmerer, D., Callaghan, M. F., Hopkins, A., Kosciessa, J., Betts, M., Cardenas-Blanco, A., Kanowski, M., Weiskopf, N., Dayan, P., Dolan, R. J., & Düzel, E. (2018). Locus coeruleus integrity in old age is selectively related to memories linked with salient negative events. Proceedings of the National Academy of Sciences of the United States of America, 115, 2228–2233. https://doi.org/10.1073/pnas.1712268115

44. Hanseeuw, B. J., Betensky, R. A., Jacobs, H. I. L., Schultz, A. P., Sepulcre, J., Becker, J. A., Cosio, D. M. O., Farrell, M., Quiroz, Y. T., Mormino, E. C., Buckley, R. F., Papp, K. V., Amariglio, R. A., Dewachter, I., Ivanoiu, A., Huijbers, W., Hedden, T., Marshall, G. A., Chhatwal, J. P., … Johnson, K. (2019). Association of amyloid and tau with cognition in preclinical Alzheimer disease. JAMA Neurology, Advance online publication. https://doi.org/10.1001/jamaneurol.2019.1424

45. Heneka, M. T., Nadrigny, F., Regen, T., Martinez-Hernandez, A., Dumitrescu-Ozimek, L., Terwel, D., Jardanhazi-Kurutz, D., Walter, J., Kirchhoff, F., Hanisch, U. K., & Kummer, M. P. (2010). Locus ceruleus controls Alzheimer’s disease pathology by modulating microglial functions through norepinephrine. Proceedings of the National Academy of Sciences of the United States of America, 107(13), 6058–6063. https://doi.org/10.1073/pnas.0909586107

46. Jacobs, H. I. L., Becker, A., Sperling, R. A., Guzman-Velez, E., Baena, A., Uquillas, F. d’Oleire, Artola, A., Pardilla-Delgado, E., Gatchel, J. R., Reiman, E. M., Johnson, K. A., Lopera, F., & Quiroz, Y. T. (2019). P2-425: Locus coeruleus intensity is associated with early amyloid and tau pathology in preclinical autosomal dominant Alzheimer’s disease. Alzheimer’s & Dementia, 15, P774–P775. https://doi.org/10.1016/j.jalz.2019.06.2832

47. Jacobs, H. I. L., Riphagen, J. M., Ramakers, I. H. G. B., & Verhey, F. R. J. (2019). Alzheimer’s disease pathology: Pathways between central norepinephrine activity, memory, and neuropsychiatric symptoms. Molecular Psychiatry, Advance online publication. https://doi.org/10.1038/s41380-019-0437-x

48. Jagust, W. (2018). Imaging the evolution and pathophysiology of Alzheimer disease. Nature Reviews Neuroscience, 19, 687–700. https://doi.org/10.1038/s41583-018-0067-3

49. Janssen, J. C., Hall, M., Fox, N. C., Harvey, R. J., Beck, J., Dickinson, A., Campbell, T., Collinge, J., Lantos, P. L., Cipolotti, L., Stevens, J. M., & Rossor, M. N. (2000). Alzheimer’s disease due to an intronic presenilin-1 (PSEN1 intron 4) mutation. A clinicopathological study. Brain, 123(5), 894–907. https://doi.org/10.1093/brain/123.5.894

50. Jessen, F., Spottke, A., Boecker, H., Brosseron, F., Buerger, K., Catak, C., Fliessbach, K., Franke, C., Fuentes, M., Heneka, M. T., Janowitz, D., Kilimann, I., Laske, C., Menne, F., Nestor, P., Peters, O., Priller, J., Pross, V., Ramirez, A., … Düzel, E. (2018). Design and first baseline data of the DZNE multicenter observational study on predementia Alzheimer’s disease (DELCODE). Alzheimer’s Research & Therapy, 10, 15. https://doi.org/10.1186/s13195-017-0314-2

51. Kelly, S. C., He, B., Perez, S. E., Ginsberg, S. D., Mufson, E. J., & Counts, S. E. (2017). Locus coeruleus cellular and molecular pathology during the progression of Alzheimer’s disease. Acta Neuropathologica Communications, 5(1), 8. https://doi.org/10.1186/s40478-017-0411-2

52. Keren, N. I., Lozar, C. T., Harris, K. C., Morgan, P. S., & Eckert, M. A. (2009). In vivo mapping of the human locus coeruleus. NeuroImage, 47(4), 1261–1267. https://doi.org/10.1016/j.neuroimage.2009.06.012

53. Keren, N. I., Taheri, S., Vazey, E. M., Morgan, P. S., Granholm, A. C. E., Aston-Jones, G., & Eckert, M. A. (2015). Histologic validation of locus coeruleus MRI contrast in post- mortem tissue. NeuroImage, 113, 235–245. https://doi.org/10.1016/j.neuroimage.2015.03.020

54. Keresztes, A., Bender, A. R., Bodammer, N. C., Lindenberger, U., Shing, Y. L., & Werkle- Bergner, M. (2017). Hippocampal maturity promotes memory distinctiveness in childhood and adolescence. Proceedings of the National Academy of Sciences of the United States of America, 114(34), 9212–9217. https://doi.org/10.1073/pnas.1710654114

55. Klein, A., Andersson, J., Ardekani, B. A., Ashburner, J., Avants, B., Chiang, M.-C., Christensen, G. E., Collins, D. L., Gee, J., Hellier, P., Song, J. H., Jenkinson, M., Lepage, C., Rueckert, D., Thompson, P., Vercauteren, T., Woods, R. P., Mann, J. J., & Parsey, R. V. (2009). Evaluation of 14 nonlinear deformation algorithms applied to human brain MRI registration. NeuroImage, 46(3), 786–802. https://doi.org/10.1016/J.NEUROIMAGE.2008.12.037

56. Krishnan, A., Williams, L. J., McIntosh, A. R., & Abdi, H. (2011). Partial Least Squares (PLS) methods for neuroimaging: A tutorial and review. NeuroImage, 56(2), 455–475. https://doi.org/10.1016/j.neuroimage.2010.07.034

57. La Joie, R., Visani, A. V., Baker, S. L., Brown, J. A., Bourakova, V., Cha, J., Chaudhary, K., Edwards, L., Iaccarino, L., Janabi, M., Lesman-Segev, O. H., Miller, Z. A., Perry, D. C., O’Neil, J. P., Pham, J., Rojas, J. C., Rosen, H. J., Seeley, W. W., Tsai, R. M., … Rabinovici, G. D. (2020). Prospective longitudinal atrophy in Alzheimer’s disease correlates with the intensity and topography of baseline tau-PET. Science Translational Medicine, 12(524), eaau5732. https://doi.org/10.1126/scitranslmed.aau5732

58. Lee, S., Viqar, F., Zimmerman, M. E., Narkhede, A., Tosto, G., Benzinger, T. L. S., Marcus, D. S., Fagan, A. M., Goate, A., Fox, N. C., Cairns, N. J., Holtzman, D. M., Buckles, V., Ghetti, B., McDade, E., Martins, R. N., Saykin, A. J., Masters, C. L., Ringman, J. M., … Network, for the D. I. A. (2016). White matter hyperintensities are a core feature of Alzheimer’s disease: Evidence from the dominantly inherited Alzheimer network. Annals of Neurology, 79(6), 929–939. https://doi.org/10.1002/ana.24647

59. Lee, T.-H., Greening, S. G., Ueno, T., Clewett, D., Ponzio, A., Sakaki, M., & Mather, M. (2018). Arousal increases neural gain via the locus coeruleus–noradrenaline system in younger adults but not in older adults. Nature Human Behaviour, 2(5), 356–366. https://doi.org/10.1038/s41562-018-0344-1

60. Lindenberger, U., von Oertzen, T., Ghisletta, P., & Hertzog, C. (2011). Cross-sectional age variance extraction: What’s change got to do with it? Psychology and Aging, 26(1), 34– 47. https://doi.org/10.1037/a0020525

61. Liu, K. Y., Acosta-Cabronero, J., Cardenas-Blanco, A., Loane, C., Berry, A. J., Betts, M. J., Kievit, R. A., Henson, R. N., Düzel, E., Howard, R., & Hämmerer, D. (2019). In vivo visualization of age-related differences in the locus coeruleus. Neurobiology of Aging, 74, 101–111. https://doi.org/10.1016/j.neurobiolaging.2018.10.014

62. Liu, K. Y., Kievit, R. A., Tsvetanov, K. A., Betts, M. J., Düzel, E., Rowe, J. B., Howard, R., & Hämmerer, D. (2020). Noradrenergic-dependent functions are associated with age- related locus coeruleus signal intensity differences. Nature Communications, 11(1), 1712. https://doi.org/10.1038/s41467-020-15410-w

63. Liu, K. Y., Marijatta, F., Hämmerer, D., Acosta-Cabronero, J., Düzel, E., & Howard, R. J. (2017). Magnetic resonance imaging of the human locus coeruleus: A systematic review. Neuroscience and Biobehavioral Reviews, 83, 325–355. https://doi.org/10.1016/j.neubiorev.2017.10.023

64. Lyness, S. A., Zarow, C., & Chui, H. C. (2003). Neuron loss in key cholinergic and aminergic nuclei in Alzheimer disease: A meta-analysis. In Neurobiology of Aging (Vol. 24, Issue 1, pp. 1–23). Elsevier. https://doi.org/10.1016/S0197-4580(02)00057-X

65. Mäki-Marttunen, V., & Espeseth, T. (2020). Uncovering the locus coeruleus: comparison of localization methods for functional analysis. NeuroImage, 117409. https://doi.org/10.1016/j.neuroimage.2020.117409

66. Makropoulos, A., Robinson, E. C., Schuh, A., Wright, R., Fitzgibbon, S., Bozek, J., Counsell, S. J., Steinweg, J., Vecchiato, K., Passerat-Palmbach, J., Lenz, G., Mortari, F., Tenev, T., Duff, E. P., Bastiani, M., Cordero-Grande, L., Hughes, E., Tusor, N., Tournier, J. D., … Rueckert, D. (2018). The developing human connectome project: A minimal processing pipeline for neonatal cortical surface reconstruction. NeuroImage, 173, 88–112. https://doi.org/10.1016/j.neuroimage.2018.01.054

67. Marien, M. R., Colpaert, F. C., & Rosenquist, A. C. (2004). Noradrenergic mechanisms in neurodegenerative diseases: A theory. Brain Research Reviews, 45(1), 38–78. https://doi.org/10.1016/j.brainresrev.2004.02.002

68. Mather, M., Clewett, D., Sakaki, M., & Harley, C. W. (2016). Norepinephrine ignites local hotspots of neuronal excitation: How arousal amplifies selectivity in perception and memory. Behavioral and Brain Sciences, 39, e200. https://doi.org/10.1017/S0140525X15000667

69. Mather, M., & Harley, C. W. (2016). The locus coeruleus: Essential for maintaining cognitive function and the aging brain. Trends in Cognitive Sciences, 20(3), 214–226. https://doi.org/10.1016/j.tics.2016.01.001

70. McIntosh, A. R., & Lobaugh, N. J. (2004). Partial least squares analysis of neuroimaging data: Applications and advances. NeuroImage, 23(SUPPL. 1). https://doi.org/10.1016/j.neuroimage.2004.07.020

71. Miyoshi, F., Ogawa, T., Kitao, S. I., Kitayama, M., Shinohara, Y., Takasugi, M., Fujii, S., & Kaminou, T. (2013). Evaluation of Parkinson disease and Alzheimer disease with the use of neuromelanin MR imaging and123I-metaiodobenzylguanidine scintigraphy. American Journal of Neuroradiology, 34(11), 2113–2118. https://doi.org/10.3174/ajnr.A3567

72. Montine, T. J., Phelps, C. H., Beach, T. G., Bigio, E. H., Cairns, N. J., Dickson, D. W., Duyckaerts, C., Frosch, M. P., Masliah, E., Mirra, S. S., Nelson, P. T., Schneider, J. A., Thal, D. R., Trojanowski, J. Q., Vinters, H. V, & Hyman, B. T. (2012). National Institute on Aging-Alzheimer’s Association guidelines for the neuropathologic assessment of Alzheimer’s disease: a practical approach. Acta Neuropathologica, 123(1), 1–11. https://doi.org/10.1007/s00401-011-0910-3

73. Moradi, E., Hallikainen, I., Hänninen, T., & Tohka, J. (2017). Rey’s Auditory Verbal Learning Test scores can be predicted from whole brain MRI in Alzheimer’s disease. NeuroImage: Clinical, 13, 415–427. https://doi.org/10.1016/j.nicl.2016.12.011

74. Muehlroth, B. E., Sander, M. C., Fandakova, Y., Grandy, T. H., Rasch, B., Lee Shing, Y., & Werkle-Bergner, M. (2020). Memory quality modulates the effect of aging on memory consolidation during sleep: Reduced maintenance but intact gain. NeuroImage, 209. https://doi.org/10.1016/j.neuroimage.2019.116490

75. Murrell, J., Ghetti, B., Cochran, E., Macias-Islas, M. A., Medina, L., Varpetian, A., Cummings, J. L., Mendez, M. F., Kawas, C., Chui, H., & Ringman, J. M. (2006). The A431E mutation in PSEN1 causing Familial Alzheimer’s Disease originating in Jalisco State, Mexico: An additional fifteen families. Neurogenetics, 7(4), 277–279. https://doi.org/10.1007/s10048-006-0053-1

76. Nakane, T., Nihashi, T., Kawai, H., & Naganawa, S. (2008). Visualization of neuromelanin in the Substantia nigra and locus ceruleus at 1.5T using a 3D-gradient echo sequence with magnetization transfer contrast. Magnetic Resonance in Medical Sciences : MRMS : An Official Journal of Japan Society of Magnetic Resonance in Medicine, 7(4), 205–210. http://www.ncbi.nlm.nih.gov/pubmed/19110515

77. Nasreddine, Z. S., Phillips, N. A., Bédirian, V., Charbonneau, S., Whitehead, V., Collin, I., Cummings, J. L., & Chertkow, H. (2005). The Montreal Cognitive Assessment, MoCA: A brief screening tool for mild cognitive impairment. Journal of the American Geriatrics Society, 53(4), 695–699. https://doi.org/10.1111/j.1532-5415.2005.53221.x

78. O’Bryant, S. E., Waring, S. C., Cullum, C. M., Hall, J., Lacritz, L., Massman, P. J., Lupo, P. J., Reisch, J. S., & Doody, R. (2008). Staging dementia using clinical dementia rating scale sum of boxes scores: A Texas Alzheimer’s research consortium study. Archives of Neurology, 65(8), 1091–1095. https://doi.org/10.1001/archneur.65.8.1091

79. O’Dell, T. J., Connor, S. A., Guglietta, R., & Nguyen, P. V. (2015). β-Adrenergic receptor signaling and modulation of long-term potentiation in the mammalian hippocampus. Learning & Memory, 22(9), 461–471. https://doi.org/10.1101/lm.031088.113

80. Parker, J., Mozaffar, T., Messmore, A., Deignan, J. L., Kimonis, V. E., & Ringman, J. M. (2019). Homozygosity for the A431E mutation in PSEN1 presenting with a relatively aggressive phenotype. Neuroscience Letters, 699, 195–198. https://doi.org/10.1016/j.neulet.2019.01.047

81. Penny, W., Friston, K., Ashburner, J., Kiebel, S., & Nichols, T. (2007). Statistical Parametric Mapping: The Analysis of Functional Brain Images. In Statistical Parametric Mapping: The Analysis of Functional Brain Images. Elsevier Ltd. https://doi.org/10.1016/B978-0-12-372560-8.X5000-1

82. Poe, G. R., Foote, S., Eschenko, O., Johansen, J. P., Bouret, S., Aston-Jones, G., Harley, C. W., Manahan-Vaughan, D., Weinshenker, D., Valentino, R., Berridge, C., Chandler, D. J., Waterhouse, B., & Sara, S. J. (2020). Locus coeruleus: a new look at the blue spot. Nature Reviews Neuroscience, 21(11), 644–659. https://doi.org/10.1038/s41583-020-0360-9

83. Possin, K. L., Laluz, V. R., Alcantar, O. Z., Miller, B. L., & Kramer, J. H. (2011). Distinct neuroanatomical substrates and cognitive mechanisms of figure copy performance in Alzheimer’s disease and behavioral variant frontotemporal dementia. Neuropsychologia, 49(1), 43–48. https://doi.org/10.1016/j.neuropsychologia.2010.10.026

84. Priovoulos, N., Jacobs, H. I. L., Ivanov, D., Uludag, K., Verhey, F. R. J., & Poser, B. A. (2017). High-resolution in vivo imaging of human locus coeruleus by magnetization transfer MRI at 3T and 7T. NeuroImage, 168, 127–136. https://doi.org/10.1016/j.neuroimage.2017.07.045

85. Ringman, J. M. (2005). What the study of persons at risk for familial Alzheimer’s disease can tell us about the earliest stages of the disorder: A review. Journal of Geriatric Psychiatry and Neurology, 18(4), 228–233. https://doi.org/10.1177/0891988705281878

86. Ringman, J. M., Monsell, S., Ng, D. W., Zhou, Y., Nguyen, A., Coppola, G., Van Berlo, V., Mendez, M. F., Tung, S., Weintraub, S., Mesulam, M.-M., Bigio, E. H., Gitelman, D. R., Fisher-Hubbard, A. O., Albin, R. L., & Vinters, H. V. (2016). Neuropathology of Autosomal Dominant Alzheimer Disease in the National Alzheimer Coordinating Center Database. Journal of Neuropathology & Experimental Neurology, 75(3), 284–290. https://doi.org/10.1093/jnen/nlv028

87. Rohlfing, T., Brandt, R., Menzel, R., & Maurer, C. R. (2004). Evaluation of atlas selection strategies for atlas-based image segmentation with application to confocal microscopy images of bee brains. NeuroImage, 21(4), 1428–1442. https://doi.org/10.1016/j.neuroimage.2003.11.010

88. Rorabaugh, J. M., Chalermpalanupap, T., Botz-Zapp, C. A., Fu, V. M., Lembeck, N. A., Cohen, R. M., & Weinshenker, D. (2017). Chemogenetic locus coeruleus activation restores reversal learning in a rat model of Alzheimer’s disease. Brain, 140(11), 3023– 3038. https://doi.org/10.1093/brain/awx232

89. Rosenthal, R. (1991). Meta-Analytic Procedures for Social Research. SAGE Publications, Inc. https://doi.org/10.4135/9781412984997

90. Ryman, D. C., Acosta-Baena, N., Aisen, P. S., Bird, T., Danek, A., Fox, N. C., Goate, A., Frommelt, P., Ghetti, B., Langbaum, J. B. S., Lopera, F., Martins, R., Masters, C. L., Mayeux, R. P., McDade, E., Moreno, S., Reiman, E. M., Ringman, J. M., Salloway, S., … Bateman, R. J. (2014). Symptom onset in autosomal dominant Alzheimer disease: A systematic review and meta-analysis. Neurology, 83(3), 253–260. https://doi.org/10.1212/WNL.0000000000000596

91. Sabuncu, M. R., Yeo, B. T. T., Van Leemput, K., Fischl, B., & Golland, P. (2010). A generative model for image segmentation based on label fusion. IEEE Transactions on Medical Imaging, 29(10), 1714–1729. https://doi.org/10.1109/TMI.2010.2050897

92. Sara, S. J. (2009). The locus coeruleus and noradrenergic modulation of cognition. Nature Reviews Neuroscience, 10(3), 211–223. https://doi.org/10.1038/nrn2573

93. Sasaki, M., Shibata, E., Tohyama, K., Takahashi, J., Otsuka, K., Tsuchiya, K., Takahashi, S., Ehara, S., Terayama, Y., & Sakai, A. (2006). Neuromelanin magnetic resonance imaging of locus ceruleus and substantia nigra in Parkinson’s disease. NeuroReport, 17(11), 1215–1218. https://doi.org/10.1097/01.wnr.0000227984.84927.a7

94. Satoh, A., & Iijima, K. M. (2019). Roles of tau pathology in the locus coeruleus (LC) in age- associated pathophysiology and Alzheimer’s disease pathogenesis: Potential strategies to protect the LC against aging. Brain Research, 1702, 17–28. https://doi.org/10.1016/J.BRAINRES.2017.12.027

95. Schoenberg, M. R., Dawson, K. A., Duff, K., Patton, D., Scott, J. G., & Adams, R. L. (2006). Test performance and classification statistics for the Rey Auditory Verbal Learning Test in selected clinical samples. Archives of Clinical Neuropsychology, 21(7), 693–703. https://doi.org/10.1016/j.acn.2006.06.010

96. Snider, B. J., Norton, J., Coats, M. A., Chakraverty, S., Hou, C. E., Jervis, R., Lendon, C. L., Goate, A. M., McKeel, D. W., & Morris, J. C. (2005). Novel presenilin 1 mutation (S170F) causing Alzheimer disease with lewy bodies in the third decade of life. Archives of Neurology, 62(12), 1821–1830. https://doi.org/10.1001/archneur.62.12.1821

97. Spina, S., La Joie, R., Petersen, C., Nolan, A. L., Cuevas, D., Cosme, C., Hepker, M., Hwang, J.-H., Miller, Z. A., Huang, E. J., Karydas, A. M., Grant, H., Boxer, A. L., Gorno-Tempini, M. L., Rosen, H. J., Kramer, J. H., Miller, B. L., Seeley, W. W., Rabinovici, G. D., & Grinberg, L. T. (2021). Comorbid neuropathological diagnoses in early versus late- onset Alzheimer’s disease. Brain : A Journal of Neurology, 144(7), 2186–2198. https://doi.org/10.1093/brain/awab099

98. Stratmann, K., Heinsen, H., Korf, H. W., Del Turco, D., Ghebremedhin, E., Seidel, K., Bouzrou, M., Grinberg, L. T., Bohl, J., Wharton, S. B., Den Dunnen, W., & Rüb, U. (2016). Precortical Phase of Alzheimer’s Disease (AD)-Related Tau Cytoskeletal Pathology. Brain Pathology, 26(3), 371–386. https://doi.org/10.1111/bpa.12289

99. Sun, W., Tang, Y., Qiao, Y., Ge, X., Mather, M., Ringman, J. M., & Shi, Y. (2020). A probabilistic atlas of locus coeruleus pathways to transentorhinal cortex for connectome imaging in Alzheimer’s disease. NeuroImage, 223, 117301. https://doi.org/10.1016/j.neuroimage.2020.117301

100. Takahashi, J., Shibata, T., Sasaki, M., Kudo, M., Yanezawa, H., Obara, S., Kudo, K., Ito, K., Yamashita, F., & Terayama, Y. (2015). Detection of changes in the locus coeruleus in patients with mild cognitive impairment and Alzheimer’s disease: High-resolution fast spin-echo T1-weighted imaging. Geriatrics and Gerontology International, 15(3), 334– 340. https://doi.org/10.1111/ggi.12280

101. Takeuchi, T., Duszkiewicz, A. J., Sonneborn, A., Spooner, P. A., Yamasaki, M., Watanabe, M., Smith, C. C., Fernández, G., Deisseroth, K., Greene, R. W., & Morris, R. G. M. (2016). Locus coeruleus and dopaminergic consolidation of everyday memory. Nature, 537(7620), 357–362. https://doi.org/10.1038/nature19325

102. Tanzi, R. E., & Bertram, L. (2005). Twenty years of the Alzheimer’s disease amyloid hypothesis: A genetic perspective. In Cell (Vol. 120, Issue 4, pp. 545–555). Cell Press. https://doi.org/10.1016/j.cell.2005.02.008

103. Teng, E. L., Hasegawa, K., Homma, A., Imai, Y., Larson, E., Graves, A., Sugimoto, K., Yamaguchi, T., Sasaki, H., Chiu, D., & White, L. R. (1994). The Cognitive Abilities Screening Instrument (CASI): A Practical Test for Cross-Cultural Epidemiological Studies of Dementia. International Psychogeriatrics, 6(1), 45–58. https://doi.org/10.1017/s1041610294001602

104. Theofilas, P., Dunlop, S., Heinsen, H., & Grinberg, L. T. (2015). Turning on the light within: Subcortical nuclei of the isodentritic core and their role in Alzheimer’s disease pathogenesis. Journal of Alzheimer’s Disease, 46(1), 17–34. https://doi.org/10.3233/JAD-142682

105. Theofilas, P., Ehrenberg, A. J., Dunlop, S., Di Lorenzo Alho, A. T., Nguy, A., Leite, R. E. P., Rodriguez, R. D., Mejia, M. B., Suemoto, C. K., Ferretti-Rebustini, R. E. D. L., Polichiso, L., Nascimento, C. F., Seeley, W. W., Nitrini, R., Pasqualucci, C. A., Jacob Filho, W., Rueb, U., Neuhaus, J., Heinsen, H., & Grinberg, L. T. (2017). Locus coeruleus volume and cell population changes during Alzheimer’s disease progression: A stereological study in human postmortem brains with potential implication for early- stage biomarker discovery. Alzheimer’s and Dementia, 13(3), 236–246. https://doi.org/10.1016/j.jalz.2016.06.2362

106. Tombaugh, T. (2004). Trail Making Test A and B: Normative data stratified by age and education. Archives of Clinical Neuropsychology, 19(2), 203–214. https://doi.org/10.1016/S0887-6177(03)00039-8

107. Tona, K. D., Keuken, M. C., de Rover, M., Lakke, E., Forstmann, B. U., Nieuwenhuis, S., & van Osch, M. J. P. (2017). In vivo visualization of the locus coeruleus in humans: Quantifying the test–retest reliability. Brain Structure and Function, 222(9), 4203–4217. https://doi.org/10.1007/s00429-017-1464-5

108. Uematsu, A., Tan, B. Z., Ycu, E. A., Cuevas, J. S., Koivumaa, J., Junyent, F., Kremer, E. J., Witten, I. B., Deisseroth, K., & Johansen, J. P. (2017). Modular organization of the brainstem noradrenaline system coordinates opposing learning states. Nature Neuroscience, 20(11), 1602–1611. https://doi.org/10.1038/nn.4642

109. Van Cauwenberghe, C., Van Broeckhoven, C., & Sleegers, K. (2016). The genetic landscape of Alzheimer disease: Clinical implications and perspectives. In Genetics in Medicine (Vol. 18, Issue 5, pp. 421–430). Nature Publishing Group. https://doi.org/10.1038/gim.2015.117

110. Wagatsuma, A., Okuyama, T., Sun, C., Smith, L. M., Abe, K., & Tonegawa, S. (2018). Locus coeruleus input to hippocampal CA3 drives single-trial learning of a novel context. Proceedings of the National Academy of Sciences of the United States of America, 115(2), E310–E316. https://doi.org/10.1073/pnas.1714082115

111. Wang, H., Suh, J. W., Das, S. R., Pluta, J. B., Craige, C., & Yushkevich, P. A. (2013). Multi- atlas segmentation with joint label fusion. IEEE Transactions on Pattern Analysis and Machine Intelligence, 35(3), 611–623. https://doi.org/10.1109/TPAMI.2012.143

112. Wechsler, D. (1981). WAIS-R manual: Wechsler Adult Intelligence Scale-Revised. Psychological Corporation.

113. Wechsler, D. (1997). WAIS-III : administration and scoring manual : Wechsler Adult Intelligence Scale (3rd ed.). Psychological Corporation.

114. Weinshenker, D. (2018). Long road to ruin: Noradrenergic dysfunction in neurodegenerative disease. Trends in Neurosciences, 41(4), 211–223. https://doi.org/10.1016/j.tins.2018.01.010

115. World Health Organization. (2004). ICD-10 : international statistical classification of diseases and related health problems : tenth revision (2nd ed). World Health Organization.

116. Ye, R., Rua, C., O’Callaghan, C., Jones, P. S., Hezemans, F. H., Kaalund, S. S., Tsvetanov, K. A., Rodgers, C. T., Williams, G., Passamonti, L., & Rowe, J. B. (2021). An in vivo probabilistic atlas of the human locus coeruleus at ultra-high field. NeuroImage, 225, 117487. https://doi.org/10.1016/j.neuroimage.2020.117487

117. Yescas, P., Huertas-Vazquez, A., Villarreal-Molina, M. T., Rasmussen, A., Tusié-Luna, M. T., López, M., Canizales-Quinteros, S., & Alonso, M. E. (2006). Founder effect for the Ala431Glu mutation of the presenilin 1 gene causing early-onset Alzheimer’s disease in Mexican families. Neurogenetics, 7(3), 195–200. https://doi.org/10.1007/s10048-006-0043-3

